# Campus-based genomic surveillance uncovers early emergence of a future dominant A(H3N2) influenza clade

**DOI:** 10.1101/2025.06.13.25329559

**Authors:** Matthew Scotch, Temitope O.C. Faleye, Jillian M. Wright, Sarah Finnerty, Rolf U. Halden, Arvind Varsani

## Abstract

We conducted genomic surveillance of seasonal influenza during the 2022-2023 northern hemisphere flu season on a large university setting in Southwest Arizona USA to understand the diversity, evolution, and spread within a local environment and how it relates to national data.

Through high-throughput sequencing and bioinformatics, we identified 100 positive samples (19%) from 516 clinical swabs collected at the student health clinic. We observed a dominance of subtype A(H3N2) which was consistent nationally for the 2022-2023 season. However, we found stark differences when examining subtype-specific H3 clades, which included an early dominance of clade 2a.3a.1 variants contrasting from country-level data in which 2b variants were most abundant. These variants might have contributed to the early seasonal peak on campus which lagged national trends by one month.

We used phylodynamics to understand the timing, source, and impact of clade-specific introductions on campus and observed introductions of 2b variants from North America, Europe, and Asia in early 2022 which possibly contributed to its later-season dominance on campus towards the end of 2022. We also observed the impact of 2b variants in our Bayesian epidemiological model, as its its emergence and rapid rise coincided with the peak of infection on campus.

We found several highly prevalent H3 mutations in known epitope sites that have been observed in multiple 3c.2a clades. In particular, we note the presence of N96S (N=57, 63%) which is a defining mutation of 2a.3 and 2a.3a.1 variants and has been shown to create a new potential N-glycosylation site in the globular head. We estimated vaccine effectiveness via an H3 epitope model with a range of 0.13–0.48 which overlaps with estimates for that year. Taken together, the abundance of antigenic drift mutations, in addition to our identification of numerous sequons found within HA1 (globular head) with high glycosylation potential likely contributed to moderate vaccine effectiveness on campus for that season.

As 2a.3a.1 variants became nearly the exclusive H3 clade nationally in 2023-2024 as well as 2024-2025, our identification of their dominance on campus highlights the importance of monitoring local settings as potential early examples for national and influenza trajectories. By using high-throughput sequencing and multiple bioinformatics methods, we show the importance of genomic epidemiology in semi-closed, highly-dense university settings and its potential for early insight of seasonal influenza diversity at a national scale.

## Introduction

Genomic epidemiology has transformed influenza surveillance by enabling real-time tracking of viral evolution, diversity, and spread [1, 2]. This approach informs vaccine strain selection, antiviral resistance monitoring, and outbreak response. University campuses are ideal for such studies due to their high population density, centralized health services, and social connectivity, which facilitate transmission and structured sampling.

Influenza A viruses (IAVs), responsible for both seasonal epidemics and pandemics, are classified by the antigenic properties of their surface glycoproteins, hemagglutinin (HA) and neuraminidase (NA), which are major targets of host immunity. These proteins evolve rapidly through antigenic drift, complicating vaccine effectiveness and necessitating continual surveillance [3, 4]. The exceptional genetic diversity, rapid mutation rate, and zoonotic potential of IAVs make them a persistent public health challenge [5–7].

Although influenza activity has rebounded following the COVID-19 pandemic, only a few genomic epidemiology studies have focused on university settings for understanding evolution and spread. Past efforts have included students in broader populations [8, 9] or focused on shorter time frames [10], but there remains a gap in understanding how influenza evolves in densely-populated campus environments. To address this, we conducted genomic epidemiology of influenza viruses at Arizona State University (ASU) during the 2022–2023 season, which featured an early surge in cases and a rapid post-holiday decline. As one of the largest U.S. universities [11], ASU offers a unique opportunity to assess intra-campus transmission, viral diversity, and evolutionary dynamics in a semi-closed population. By comparing these findings with national patterns, we aim to inform surveillance strategies and public health preparedness for other high-risk, congregate settings.

## Materials and methods

### Sample processing and molecular testing

As part of an on-going surveillance study, we analyzed nasopharyngeal (NP) or mid-turbinate swabs from student health services on the Tempe campus of Arizona State University (USA) during the 2022–2023 North American influenza season. The clinic staff tested students via a lateral flow immunoassay as part of routine medical care who presented with influenza-like illness (ILI). They labeled the collection tubes with the result (either influenza A, B, A/B, or negative), and provided the date of sample collection. They did not provide any patient identifiable information.

At our laboratory, we froze samples at –80*^◦^*C or immediately subjected 140 *µ*l to RNA extraction with the QIAamp Viral RNA kit according to the manufacturer’s specifications (Qiagen, Inc., Venlo, Netherlands). We performed cDNA synthesis on the extracted RNA using the universal influenza A/B UniFlu primer (5’–IAGCARAAGC–3’) described by Zhao *et al.* [12] and the SuperScript IV first strand cDNA synthesis kit (Thermo Fisher, Waltham, MA, USA) following the manufacturer’s instructions. We stored the cDNA at –20*^◦^*C and used it for downstream assays.

We performed real-time PCR to confirm detection of influenza A and B nucleic acid using the SsoAdvanced Universal Probes Supermix (Bio-Rad, Hercules, CA, USA) following the manufacturer’s instructions. For amplification, we used a QuantStudio 5 thermocycler (Thermo Fisher, Waltham, MA, USA) with reaction settings that include an initial denaturation at 95*^◦^*C for 3 minutes, followed by 40 cycles of 95*^◦^*C for 15 seconds and 60*^◦^*C for 30 seconds during which time we recorded fluorescence data.

For confirmed positive samples, we used the multiplex assay by Zhou *et al.* [13], and as previously described by [14], for amplification of the hemagglutinin (HA and neuraminidase (NA) coding segments on a BioRad C1000 thermal cycler (BioRad, Hercules, CA, USA). We used the cDNA along with the forward and reverse primers from Table S1 and Phusion-plus master mix (Thermo Fisher, Waltham, MA, USA) with the following thermal cycling conditions; 94*^◦^*C for 2 minutes, 5 cycles of 94*^◦^*C for 30 seconds, 45*^◦^*C for seconds, and 68*^◦^*C for 3 minutes; 35 cycles of 94*^◦^*C for 30 seconds, 57*^◦^*C for 30 seconds, and 68*^◦^*C for 3 minutes; followed by a final elongation step of 68*^◦^*C for 10 minutes and 12*^◦^*C until stopped. Finally, we purified the amplicons via magnetic beads, and prepared MinION long-read sequencing libraries (MinION SQK-LSK110 and EXP-NBD104, Oxford Nanopore Technologies, Oxford, UK) following the manufacturer’s instructions. We performed quality control, trimming, assembly, polishing, and variant annotation using standard pipelines (see *Supplemental methodology*). We uploaded our consensus FASTA sequences to Nextclade [15, 16] and removed, from our datasets, any sequences receiving an overall quality status of *bad*. For initial comparison, we aligned our segment-specific datasets via MAFFT [17] and used the Sequence Demarcation Tool (SDT) [18] v1.3 for sequencing similarity analysis.

### Bayesian phylogenetic and phylodynamic inference

We combined our new sequences with 2022–2023 U.S. sequences from GISAID to create segment and subtype-specific datasets (see *Supplemental methodology* ). We performed multiple sequence alignment in Geneious Prime v.2025.1.2 (Dotmatics, Boston, MA USA) using MAFFT v7.490 [17] with default parameters and visually inspected the resulting alignment to identify and remove any misclassified sequences or alignment errors.

We used TempEST v1.5.3 [19] to assess the temporal signal and identified a strong clock-like relationship between sampling time and root-to-tip divergence in both the H3 (R^2^ *>* 0.92) and N2 (R^2^ *>* 0.87) phylogenies under the best-fitting root; supporting the use of a strict molecular clock model. We used BEAST v1.10.4 [20] for Bayesian phylogenetic inference, ancestral state reconstruction, and Markov jump counting to assess influenza virus introductions into our campus (see *Supplemental methodology*).

### Bayesian epidemiological modeling

To understand the population dynamics of seasonal influenza on a large university campus, we leveraged the *PhyDyn* package as part of BEAST2 [21]. This approach allows for the simultaneous estimation of epidemiological-relevant parameters (such as R0) and coalescent-based phylogenies [21, 22]. The epidemic models in PhyDyn are implemented through ordinary differential equations (ODEs) to estimate the rates of new infections and transitions between states. We created an additional dataset of sequences from the 2022–2023 season. Here, we implemented a stratified sampling strategy to ensure representative coverage with both temporal and geographic diversity. First, we grouped sequences into weekly intervals and randomly selected up to ten sequences per week, Second, we proportionally sampled based on the availability of sequences from each of the ten Health and Human Services (HHS) regions [23]. To maximize diversity, we combined sequences sampled in both strategies and removed duplicates. In Geneious Prime v2024.0.7 (Dotmatics, Boston, MA USA) , we removed records that reflected incomplete coding regions (e.g. lacking a start and/or stop codon) and performed multiple sequence alignment via the mafft plugin v7.490 [17] with default settings.

For our H3 dataset, we utilized a two-deme SIR model adapted from Volz [24] where *I* represents our campus population while *N_r_* is an estimate of the effective population size of a global A(H3N2) community (see *Supplemental methodology* ).

### Mutation profiles, vaccine protection estimates, and glycosylation sites

We translated our HA and NA gene segments into protein sequences and checked for variants and phenotypic importance via FluServer [25] by specifying the recommended vaccine strains for each subtype (Table S2). For each query, we examined the mutations interest level (0-3), any previously reported effects or multiple position effects, and known structural interactions. For neuraminidase, we additionally considered whether there were mutations with known antiviral resistance.

As the effectiveness of influenza vaccines (VE) changes from season-to-season, we analyzed the protection of the vaccine among our translated H3 hemagglutinin sequences via pEpitope [26] v2.2 in MATLAB. Finally, we used the NetNGlyc-1.0 server [27] to predict N-glycosylation sites which considers N-X-S/T motifs where *X* can be any amino acid except proline.

### Use of Generative AI

We used an institutional license of ChatGPT 4o to generate R code for visualization of Markov jump introductions and clade frequencies (Fig. 2 and Fig. 3), lollipop graphs of amino acid mutations (Fig. 4), bar charts of median depth (Fig. S3), SNP figures (Fig. S4–S7), ANOVA calculations (Table S8), and monophyletic assessment of the MCC trees (Fig. 2 and Fig. 3). We also used it to edit python code including for visualization of the MCC trees (Figs. 2–3 and Figs. S13–S14).

**Fig 1.**
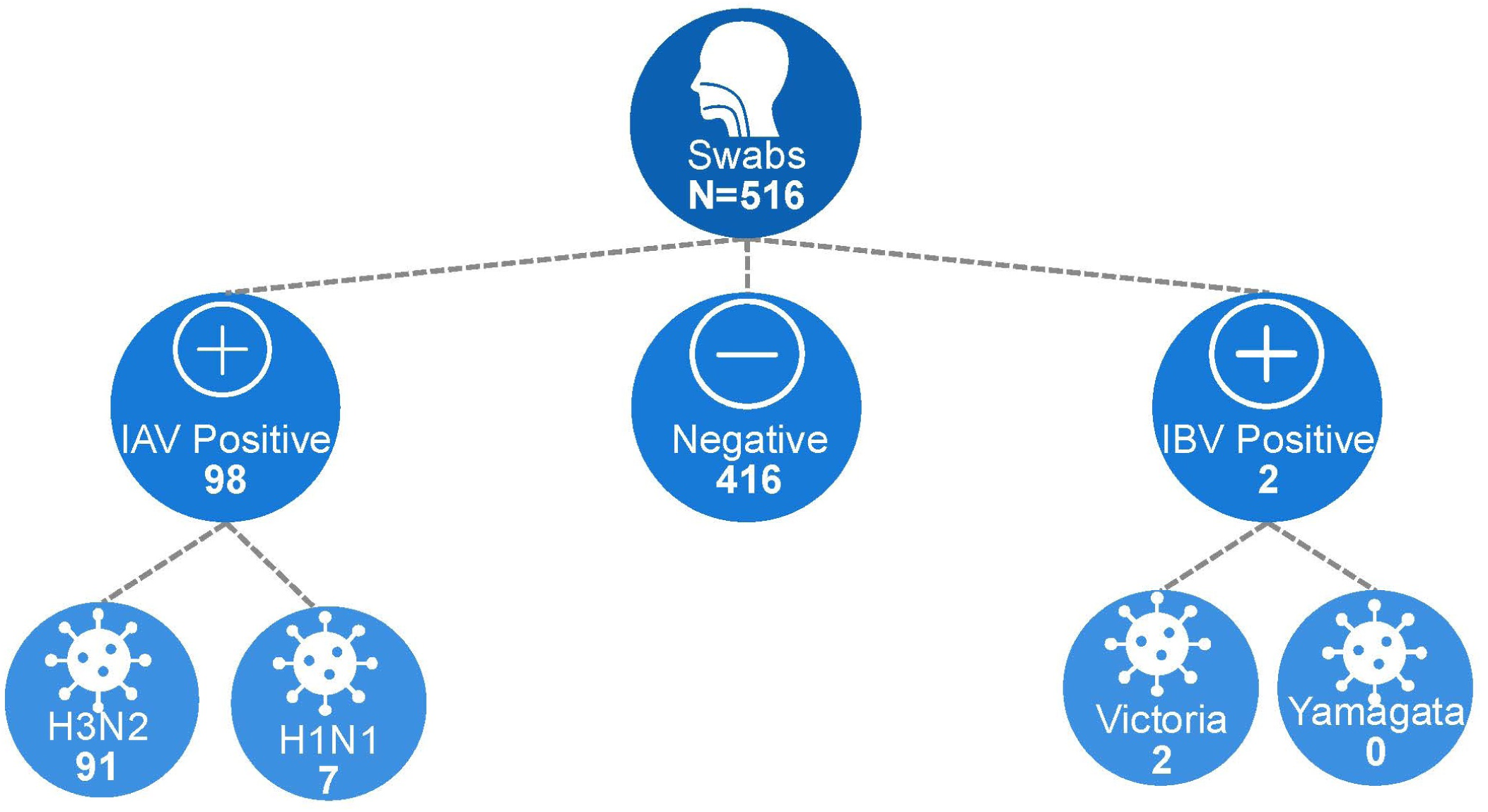
Results of our 516 clinical swabs collected at the student health clinic. Providers tested samples at the student health clinic as part of routine clinical care and labeled as ‘A’, ‘B’, ‘A/B’, or negative via a lateral flow immunoassay. We then subtyped positives via real-time PCR and pathogen sequencing. Abbreviations: IAV = Influenza A Virus; IBV = Influenza B Virus. Note: While H3 was confirmed in 91 samples, N2 was only confirmed in 76 of these; therefore, it is possible that not all H3 samples were A(H3N2).

**Fig 2.**
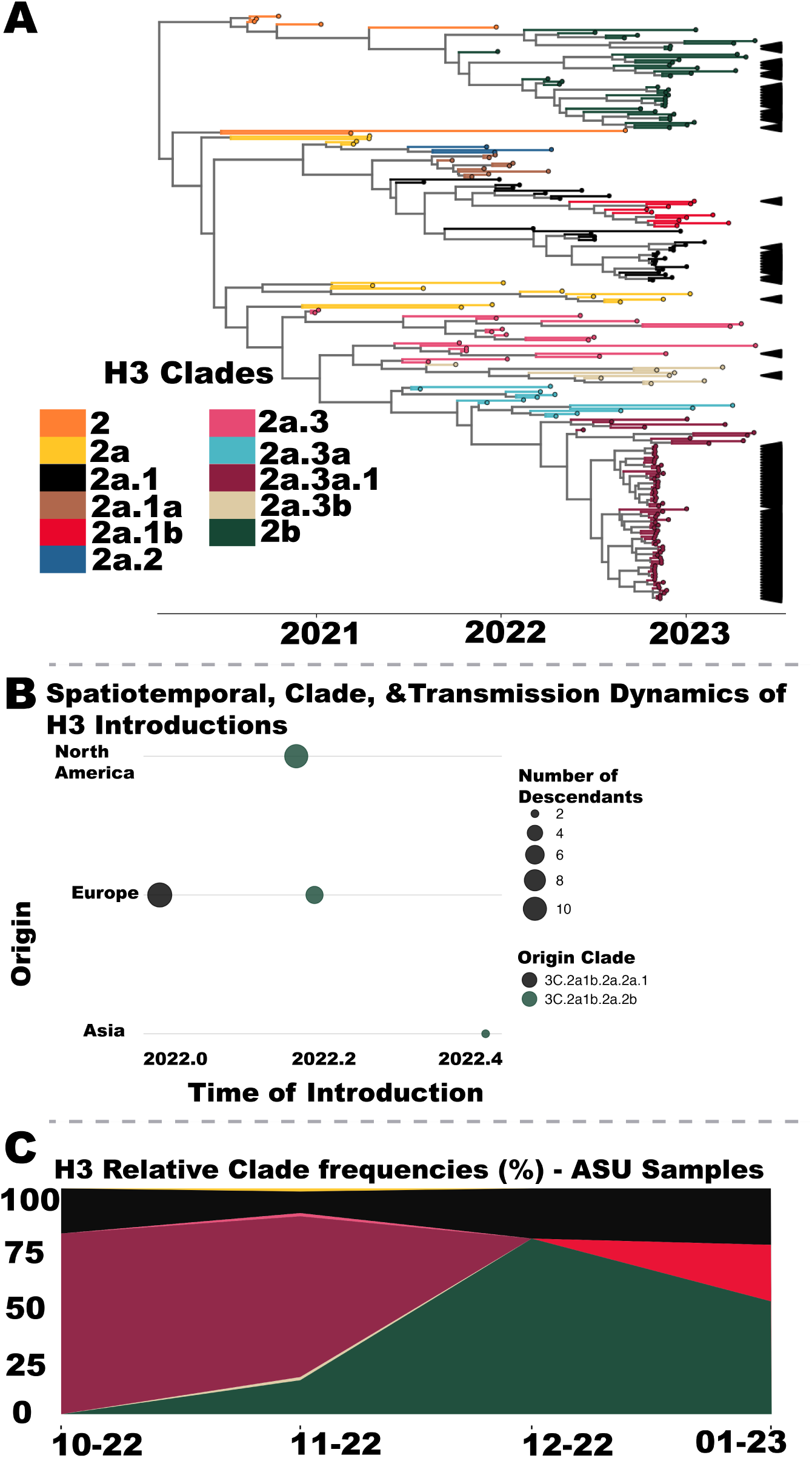
Spatiotemporal and evolutionary dynamics of H3 influenza clades circulating at ASU. In panel A, we show a sub-tree of the MCC tree from Fig. S13, pruned to include only sequences descended from the TMRCA of the ASU samples. Black arrows indicate ASU sequences (*N* = 91); all other tips represent GISAID sequences. We color-code branches by 3c.2a1b clades using abbreviated names (e.g., ‘3c.2a1b.2a.2a.3a.1’ for ‘2a.3a.1’). In panel B, we summarize the timing, geographic origin, and transmission dynamics of H3 introductions into ASU, with circle sizes reflecting the number of sampled descendants per introduction. In panel C, we display the monthly distribution of H3 clade frequencies among our university samples from October 2022 to January 2023 using the color scheme depicted in panel A. We finalized axis labels and legends in Adobe Illustrator.

**Fig 3.**
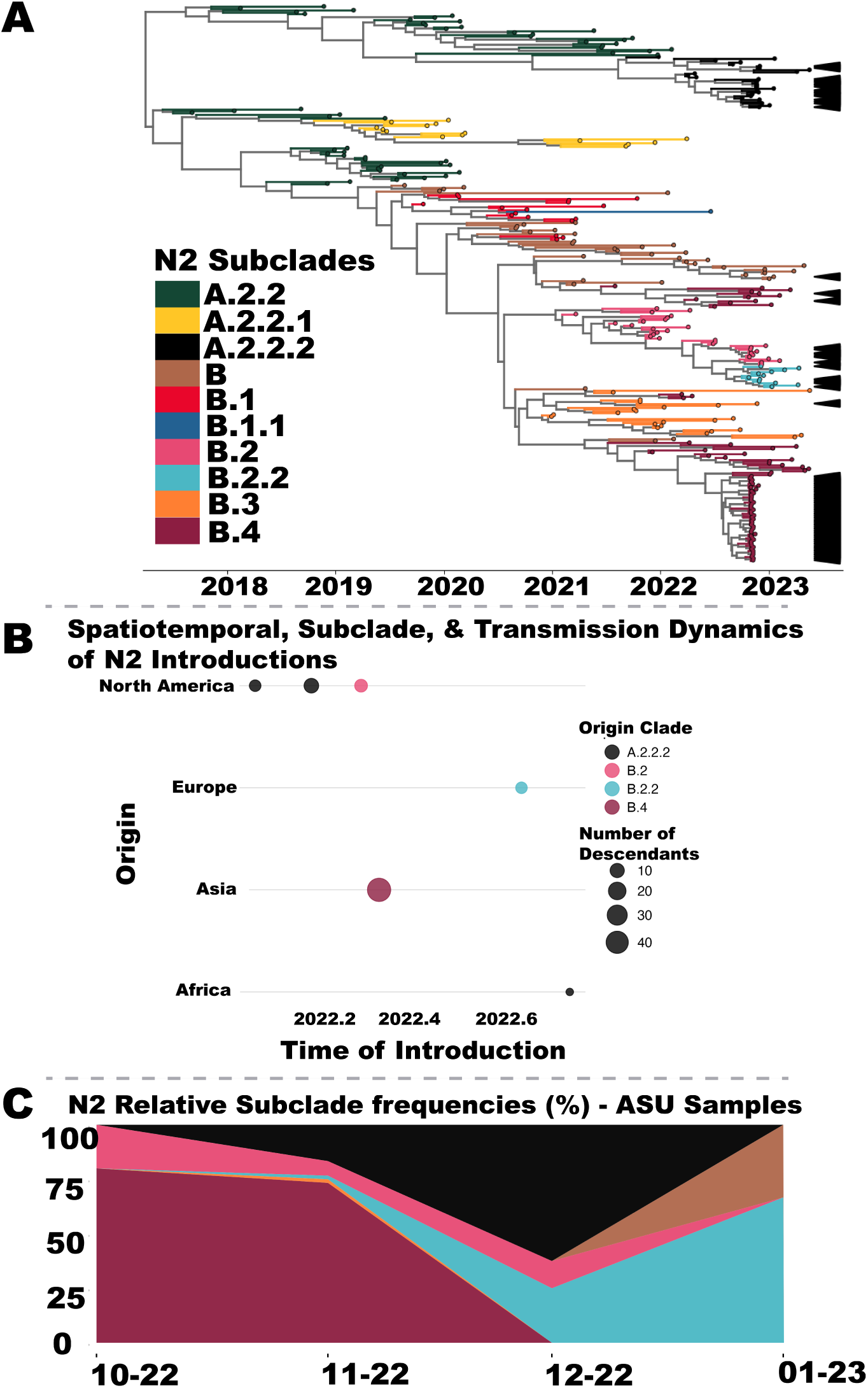
Spatiotemporal and evolutionary dynamics of N2 influenza clades circulating at ASU. In panel A, we show a sub-tree of the MCC tree from Fig. S14, pruned to include only sequences descended from the TMRCA of the ASU samples. Black arrows indicate ASU sequences (*N* = 76); all other tips represent GISAID sequences. We color-code branches by subclade using abbreviated names (e.g., ‘A.2.2.1’). In panel B, we summarize the timing, geographic origin, and transmission dynamics of H3 introductions into ASU, with circle sizes reflecting the number of sampled descendants per introduction. In panel C, we display the monthly distribution of N2 clade frequencies among our university samples from October 2022 to January 2023 using the color scheme depicted in panel A. We finalized axis labels and legends in Adobe Illustrator.

**Fig 4.**
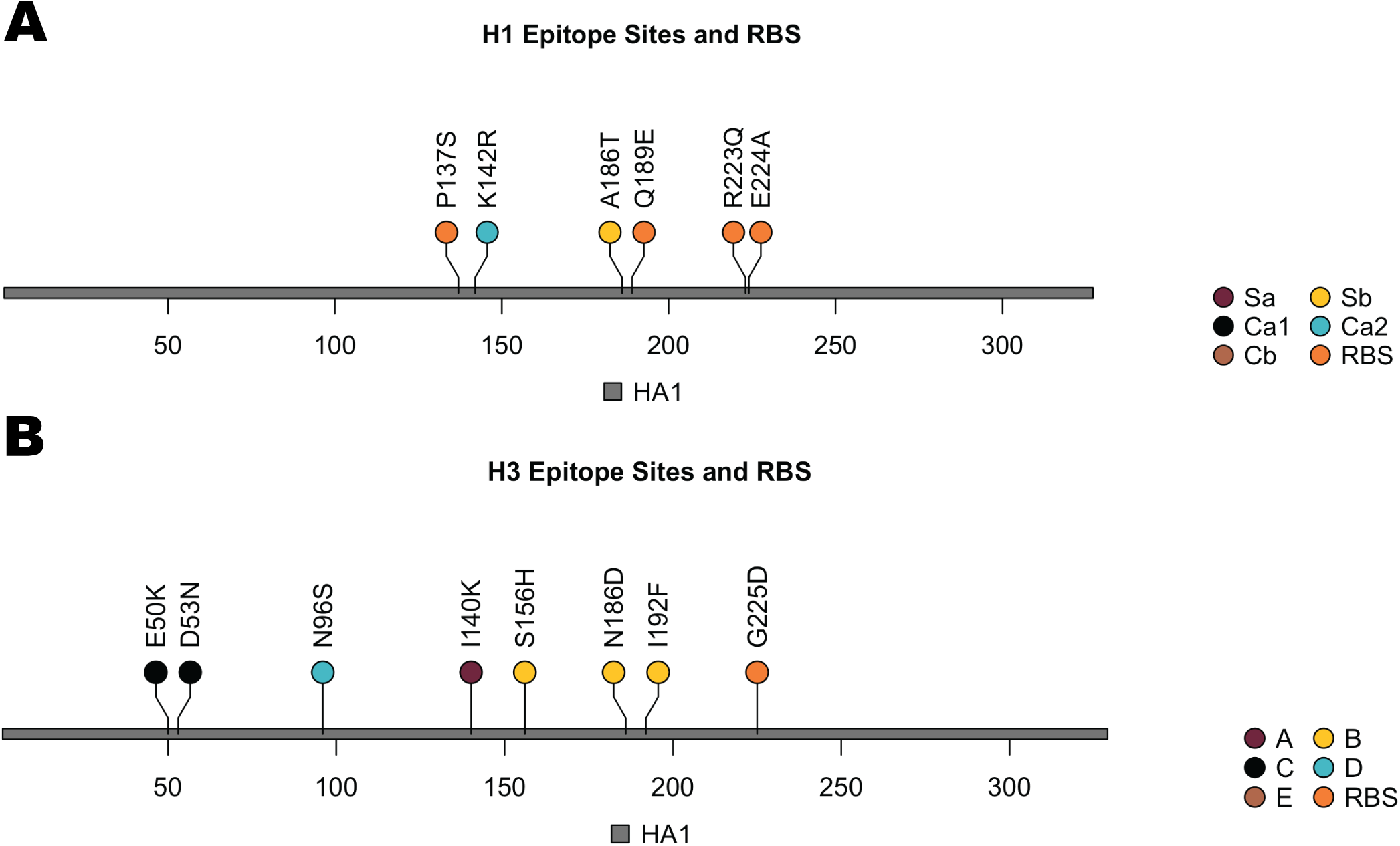
Amino acid mutations from our university influenza A virus sequences with a prevalence of *≥* 0.2 and are located in known epitope and receptor binding sites. A) Lollipop plot showing amino acid mutations for our H1 virus sequences (H1 numbering). B) Amino acid mutations for our H3 virus sequences (H3 numbering). Abbreviations: RBS - receptor binding site.

## Results

In Fig. 1, we show the final determination of the 516 nasal swab samples that we analyzed from the university’s student health clinic on the Tempe campus during the 2022–2023 season. For the 100 positives (19%), we identified 91 A(H3N2), 7 A(H1N1)pdm09, and 2 B(Victoria). Meanwhile, 416 samples tested negative. We note that while H3 was confirmed in 91 samples, N2 was only confirmed in 76 of these; therefore, it is possible (though unlikely) that not all of the 15 samples lacking an N2 sequences were A(H3N2). In Fig. S1, we show the results by week of collection starting from week 43 in 2022 to week 19 in 2023. In November, the early peak in cases on campus occurred four weeks before (44 compared to 48) the national peak (Fig. S2). The earlier season peak was likely a contributing factor to the sharp decline in cases after 2023 which resulted in almost all negative samples on campus.

For our 100 positive university samples, we generated 2,042,446 raw reads, of which 1,710,494 (83.75%) passed quality thresholds, and 1,525,574 (89.19%) of those were successfully mapped by IRMA. The median coverage depth varied across gene segments, with H3 exhibiting by far the highest mean depth (Fig. S3). All 91 H3 segments met the coverage threshold of 100, while 76 out of 87 (87%) N2 segments satisfied the criteria (TableS3). Through variant analysis, we identified synonymous and missense mutations for H3 (Fig. S4), N2 (Fig. S5), H1 (Fig. S6), and N1 (Fig. S7). For H3, we identified 2,182 variants, the majority of which were synonymous (1,290, or 59%), followed by missense mutations (892, or 41%). N2 showed a similar distribution among its 1,118 variants, with most being synonymous (802, or 74%) in comparison to missense (284, or 26%).

### Sequence similarity and clade diversity

Throughout this manuscript, we refer to clades of the HA (H3) segment using the standard ‘3c’ nomenclature (*e.g.*, clades 3C.2a1b.2b, 3C.2a2), consistent with common practice. In contrast, for the NA (N2) segment, we follow Nextstrain’s nomenclature [28] for *subclades* that designates major lineages and sub-lineages (e.g., clades A and B, subclades A.2.2, B.3, etc.).

We show strong sequence similarity for our H3 (Fig. S8) and N2 (Fig. S9) sequences, with average identity scores of 0.990 and 0.989, respectively. Fifty-six of the 91 university H3 sequences (62%) belonged to clade 2a.3a.1, 20 (22%) to 2b, and 11 (12%) to 2a.1 (Fig. S10, panel A). The remaining four sequences (4%) represented 2a, 2a.1b, 2a.3, and 2a.3b. Notably, while clade 2b was the dominant lineage nationally during the same season (accounting for over 70% of sequences at times) [29], it was a minority on campus, suggesting localized transmission dynamics that diverged from national trends. For N2 (Fig. S10, panel B), 63% of sequences (48/76) were classified as B.4, followed by 20% (15/76) as A.2.2.2, with the remainder representing B.2 (6), B.2.2 (5), and B (1).

In contrast to the dominance of A(H3N2) at both the university and national levels, A(H1N1)pdm09 was far less prevalent in our campus dataset. While A(H3N2) comprised 93% of our A/HA sequences, only 7% were H1 and in comparison to 27% nationally during the 2022–2023 season [30]. Like H3 and N2, our H1 and N1 sequences exhibited strong similarity (Fig. S11) with an average identity score of 0.988 and 0.993, respectively. Four of the seven university H1 sequences (57%) belonged to 6B.1A.5a.2a.1 while three of the sequences (43%) were 6B.1A.5a.2a (Fig. S12 panel A). For N1 (Fig. S12 panel B), three of the seven were C.5.1.1 (43%), two were C.5.3 (29%), and there was one C.2 and C.5.1 (14%).

In addition to influenza A, we also identified two influenza B/Victoria variants (2% of our samples) that were both representative of clade V1A.3a.2 which has dominated the B/Victoria landscape for several years. The sequences belonged to different subclades however, with one in C.3 and one in C.5.7.

For our Bayesian phylogenetic inference and epidemiologic modeling, we focused on subtype A(H3N2), which accounted for a significant proportion of the influenza variants circulating on campus during the study period. Its high prevalence, combined with the observed clade diversity, provided a robust foundation for downstream analyses of evolutionary trajectories and epidemiological dynamics.

### Bayesian phylogenetic and phylodynamic inference

We integrated our university-derived sequences with representative data from GISAID, resulting in a comprehensive dataset of 395 H3 and 377 N2 sequences. In Fig. 2 panel A, we present the sub-tree of the full phylogeny (Fig. S13) that includes all sequences descended from the most recent common ancestor (TMRCA) of our campus samples. For H3, we estimated the median TMRCA to be 3.22 years ago (95% highest posterior density [HPD]: 3.00–3.47 years), corresponding to approximately February 28, 2020 (95% HPD: 27 November 2019 – 18 May 2020). This predates the 2020–2021 northern hemisphere influenza season and during the pandemic period, despite historically low influenza circulation. We estimated the median rate of evolution across all H3 clades at 3.23 *×* 10*^−^*^3^ substitutions/site/year (95% HPD: 2.94 *×* 10*^−^*^3^–3.55 *×* 10*^−^*^3^).

Of the 215 sequences in the sub-tree (Fig. 2 panel A), 91 originated from the university, with representation across eleven distinct H3 clades. Six of these clades (2a.1a, 2a.1b, 2a.2, 2a.3a.1, 2a.3b, and 2b) were monophyletic, consistent with discrete introduction events. Clade 2a.3a.1 was the most common among early-season variants, while clade 2b emerged prominently later, forming a large, monophyletic cluster suggesting the occurrence of extensive local transmission on campus.

In panel B in Fig. 2, we show the timing, origin, and clades for inferred introduction events. Here, each point corresponds to a unique introduction, sized by the number of downstream descendants and colored by clade. The four distinct events highlight introductions from Asia, Europe, and North America. Introduction events linked to clade 2a.2a.1 from Europe resulted in relatively few descendants, while a North American-derived 2b introduction yielded the largest amount of observed local transmission.

We see this replacement pattern in Fig. 2 panel C, which tracks the changing distribution of clades over time. Clade 2a.3a.1 (maroon color) dominated in October and November 2022 but was quickly overtaken by clade 2b (green) in December. The 2b clade maintained its dominance until influenza activity declined sharply in January 2023, marking the end of the season on campus.

In Fig. 3 panel A, we show the sub-tree of the full N2 MCC phylogeny (Fig. S14) representing all sequences descended from the most recent common ancestor (TMRCA) of the university samples. The median TMRCA for N2 was estimated to be 6.19 years ago (95% highest posterior density [HPD]: 5.71–6.75 years), corresponding to 9 March 2017 (95% HPD: 18 August 2016 – 2 September 2017). The estimated median evolutionary rate was 2.96 *×* 10*^−^*^3^ substitutions/site/year (95% HPD: 2.63 *×* 10*^−^*^3^–3.31 *×* 10*^−^*^3^).

The MCC tree reveals two major lineages of N2, further subdivided into distinct subclades. Of the 298 sequences in the sub-tree, 76 were derived from the university and distributed across ten unique subclades. Several subclades such as A.2.2.2, B.2.2, B.3, and B.4, formed monophyletic clusters. In contrast, subclade A.2.2 appeared in multiple branches of the tree, consistent with a pattern previously reported in the literature [31].

We identified six distinct N2 introduction events in 2022 (Fig. 3 panel B), originating from Asia (B.4), North America (two separate introductions of A.2.2.2 and one of B.2), Europe (B.2.2), and Africa (A.2.2.2). The introduction of B.4 from Asia was the most impactful, generating a large local transmission chain with 46 descendant sequences (this is also reflected in Fig. 3 panel C), where B.4 dominated circulation in October and November. Two independent introductions of A.2.2.2 from North America produced 11 and 5 sequences, respectively, while the African A.2.2.2 lineage contributed two. The B.2 and B.2.2 introductions yielded 7 and 5 descendants, respectively.

In Fig. 3 panel C, we show the proportion of subclades on campus over time. Subclade B.4 (maroon color), which dominated the early season, declined sharply by December. This decline corresponded to the rise of A and then B subclades, A.2.2.2 (black) and B.2.2 (light blue), respectively. In particular, A.2.2.2 rapidly displaced B.4 in November while B.2.2 replaced A.2.2.2 and became the dominant lineages through January 2023. The rise of A.2.2.2 is primarily attributed to two separate introduction events from North America in early 2022, each leading to local transmission. A third introduction occurred at the start of the influenza season but had a much smaller transmission impact, potentially due to ongoing local circulation, consistent with observations from other viral epidemics where early introductions shape subsequent transmission dynamics. Meanwhile, the rise of B.2.2 appears to be driven in part by a single introduction from Europe in late August, shortly before the start of the season, followed by limited local expansion.

### Bayesian epidemiological modeling

Using a two-deme compartmental SIR model, we estimated a basic reproduction number (R_0_) of 1.17 (95% HPD: 1.06–1.30), which falls within the expected range, though on the lower end, for seasonal influenza outbreaks [32]. As we show in Fig. S15 panel A, the number of susceptible individuals decreased steadily over the semester as students became infected. At the start of the season in early October, the model estimated 123 infected and 132 recovered individuals. The infected population increased over the following weeks, peaking at 581 individuals in late November or early December. Notably, this epidemic peak coincided with the emergence and rapid rise of clade 2b, as shown in Fig. 2 panel C, suggesting that the observed transmission dynamics were at least partially driven by the introduction of this clade.

The dynamics of the epidemic are further reflected in Fig. S15 panel B, which shows the rate of change in each compartment. Infection rates had already begun to decline before the end of the fall semester (first dashed line), and we observed no resurgence following the start of the spring semester in January (second dashed line). These findings suggest that the epidemic had largely burned out by the start of 2023, consistent with national trends (Fig. S2) as well as a limited susceptible population that remained on campus over winter break.

### Amino acid analyses

#### Nonsynonymous mutations

For our consensus sequences, we identified 777 total amino acid mutations across our 91 H3 proteins with A/Darwin/9 as our reference strain. This equated to an average of 8.5 mutations per protein. In Table 1, we show the most prevalent mutations (*≥* 0.2) and their 3C.2a1b associated clades. In particular, G225 and N186D were found in all H3 variants sampled on campus, while I140K and E50K were present in over 80% (Fig. 4 panel B). We also observed eleven mutations that are only seen in 2b including S156H, F79V, D101E, T135A, F79I, A476T, N122S, Q356L, S137A, S54N, and S91N.

**Table 1.** H3 amino acid mutations with a prevalence of ≥ 0.2. For each mutation (H3 numbering), we show its count, proportion across our 91 variants, and associated clades (short name). We highlight in gray the mutations in known epitope regions.

| Mutation | Count | Proportion (N=91) | Associated 3C.2a1b clades |
| --- | --- | --- | --- |
| G225D | 91 | 1.00 | 2a; 2a.1; 2a.1b; 2a.3; 2a.3a.1; 2a.3b; 2b |
| N186D | 91 | 1.00 | 2a; 2a.1; 2a.1b; 2a.3; 2a.3a.1; 2a.3b; 2b |
| I140K | 77 | 0.85 | 2a.1b; 2a.3a.1; 2b |
| E50K | 76 | 0.84 | 2a.3a.1; 2b |
| D53N | 59 | 0.65 | 2a; 2a.3; 2a.3a.1; 2a.3b |
| I192F | 58 | 0.64 | 2a.3; 2a.3a.1; 2a.3b |
| N378S | 58 | 0.64 | 2a.3; 2a.3a.1; 2a.3b |
| N96S | 57 | 0.63 | 2a.3; 2a.3a.1 |
| I223V | 56 | 0.62 | 2a.3a.1 |
| V204I | 23 | 0.25 | 2a.3a.1 |
| S156H | 20 | 0.22 | 2b |
| F79V | 18 | 0.20 | 2b |

We observed 238 total N2 amino acid mutations from A/Darwin/9 across our 76 proteins for an average of 3.13 variants per protein (Table 2). This includes one instance of mutation S331R which has been showed to result in reduced susceptibility to oseltamivir and zanamivir [33].

**Table 2.** N2 amino acid mutations with a prevalence of ≥ 0.2. For each mutation, we show its count, proportion across our 76 variants.

| Mutation | Count | Proportion (N=76) |
| --- | --- | --- |
| S150H | 47 | 0.62 |
| S150R | 29 | 0.38 |
| V263I | 16 | 0.21 |
| D346S | 15 | 0.2 |
| N463D | 15 | 0.2 |
| R315S | 15 | 0.2 |
| S465N | 15 | 0.2 |

For our seven A(H1N1)pdm09 sequences, we identified 97 total H1 (Table S5, Fig. 4 panel A) and 20 total N1 (Table S6) amino acid mutations with A/Victoria/2570/2019 as the reference strain. This equated to an average of 13.9 (H1) and 2.86 (N1) mutations per protein, respectively. None of the N1 mutations have been showed to impact antiviral sensitivity (Table S6).

For our two B/Victoria variants, one HA segment had three mutations including E121G, E169K, and D184E, while the other had four mutations including E121K, A146E, E185G, and S195P (against reference strain B/Austria/1359417/2021). In regards to NA mutations, one virus sequence had A44V, G372E, and I459V, while the other had R57H, V388I, and I459V. None of the NA mutations have been linked to antiviral drug resistance.

#### Vaccine effectiveness and N-glycosylation sites

Among 25 unique translated H3 sequences from the university, we observed a variation of vaccine effectiveness estimates (0.13 - 0.48) and dominant epitope regions (Table S7). Region B was the most common dominant epitope impacted (16/25 or 64%), followed by A (5/25, or 20%) and then C (4/25 or 16%). No sequences included D or E as dominant epitopes. We observed I140K as the most commonly seen mutation for epitope A in 19 out of the 25 unique sequences (76%), N186D in all 25 sequences for epitope B, and E50K in 18 of the 25 unique sequences for epitope C (72%). We performed an ANOVA analysis in SPSS and found marginal differences in predicted VE across the dominant epitope groups, but the differences were not statistically significant (p=0.071, Table S8).

We leveraged NetNGlyc v1.0 to identify asparagine-Xaa-serine/threonine sequons and predict N-glycosylation sites within our 91 H3 sequences. In total, we observed fourteen unique sequons (for thirteen sites). Nine of the sequons were observed in all sequences, while two were observed in all but one sequence. Four sequons had an average potential *>* 0.7 including NSTA (position 24), NGTI (38), NCTL (79), and NVTM (181). In Table 3, we show the sequons with unanimous jury agreement across nine neural networks. (ordered by position on the H3 protein). Of note, NTSA (position 24), NCTL (position 79), and NGSI (position 301), were predicted as N-glycosylated and found in all sequences with high specificity. Not surprisingly, all sequons listed in Table 3 are found in the HA1 subunit (globular head).

**Table 3.** Summary of H3 sequons with glycosylation potential among our 91 H3 samples as estimated by NetNGlyc - 1.0. We include sequons with high specificity and unanimous agreement across all nine neural networks and ordered by position on the HA protein (H3 numbering).

| Position | Sequon | Jury Agreement | Count | Potential (mean) | N-Glyc Result |
| --- | --- | --- | --- | --- | --- |
| 24 | NSTA | (9/9) | 91 | 0.79 | +++ |
| 38 | NGTI | (9/9) | 90 | 0.7 | ++ |
| 38 | NGTM | (9/9) | 1 | 0.66 | ++ |
| 79 | NCTL | (9/9) | 91 | 0.74 | ++ |
| 149 | NGTS | (9/9) | 14 | 0.64 | ++ |
| 181 | NVTM | (9/9) | 90 | 0.75 | ++ |
| 262 | NSTG | (9/9) | 3 | 0.64 | ++ |
| 301 | NGSI | (9/9) | 91 | 0.68 | ++ |

## Discussion

Universities represent an important, yet, understudied environment for influenza genomic epidemiology due to their semi-closed environment, close mixing of individuals, and potential early indication of state and national-level trends. The 2022-2023 influenza season in the United States signified a near-return to pre-pandemic levels [10] as well as a historically early peak in clinical cases resulting in an early population bottleneck and end to the influenza season. The confluence of these factors make the 2022-2023 season an important time-point in university-focused influenza evolution and dynamics.

We conducted genomic surveillance of seasonal influenza viruses during the 2022-2023 northern hemisphere season on a large university setting in Southwest Arizona USA to understand the diversity, evolution, and spread within a local environment and in comparison to national and international trends. We focused on the amplification and sequencing of hemagglutinin (HA) and neuraminidase (NA) gene segments for influenza A and B seasonal viruses. We observed a dominance of influenza A and, in particular, subtype A(H3N2) (Fig. 1, Fig. S1) which was consistent with national trends for that season [30]. However, we found stark differences when examining subtype-specific H3 clades, which included an early dominance of 2a.3a.1 variants (Fig. 2) contrasting from national-level data in which 2b were the most abundant. These variants might have contributed to the early seasonal peak on campus as compared to national trends (Fig. S2). As 2a.3a.1 variants became nearly the exclusive H3 clade nationally in 2023-2024 [34] as well as 2024-2025 [35], our identification of their presence on campus highlights the importance of monitoring local settings as potential early examples for national and influenza trends.

We used phylodynamics to understand the timing, source, and impact of clade-specific introductions on campus (Fig. 2). Interestingly, we did not observe any 2a.3a.1 introductions suggesting the potential for earlier cryptic transmission [36] or as a result of insufficient sampling potentially exacerbated by the early peak on campus. We did, however, find introductions of 2b variants from North America, Europe, and Asia in early 2022 which possibly contributed to its later-season local dominance towards the end of 2022. The rise of clade 2b was also impactful in our Bayesian epidemiological model, as we observed a peak in the population of infected individuals on campus during its emergence and rapid rise (Fig. S15).

As H3 sequences reveal an important, yet incomplete explanation of seasonal influenza evolution and dynamics, we also studied N2 subclades (Fig. 3, Fig. S10) and found a similar pattern of early dominance of one group followed by a replacement by another, in B.4 and A.2.2.2 respectively. Phylogenically, both of these subclades formed monophyletic clusters on our MCC tree (Fig. 3) potentially due to introduction events, albeit from different sources. The early dominance of B.4 variants was likely influenced by a single introduction even in early 2022 from Asia which led to significant transmission in our campus environment. Meanwhile, we observed two introduction events for the A.2.2 subclade including one from Africa right around the start of the influenza season which potentially contributed to its eventual takeover of B.4 variants on campus.

We utilized Bayesian phylogenetic inference to estimate the TMRCA of our university H3 and N2 sequences and estimated the median TMRCA of H3 to be 3.22 years ago (95% highest posterior density [HPD]: 3.00–3.47 years), which predates the 2020–2021 northern hemisphere influenza season. Correspondingly, we estimated the median rate of evolution across all H3 clades at 3.23 *×* 10*^−^*^3^ substitutions/site/year (95% HPD: 2.94 *×* 10*^−^*^3^–3.55 *×* 10*^−^*^3^), consistent with some previous reports [37, 38] but slower than others [39, 40]. For N2, the estimated median evolutionary rate was 2.96 *×* 10*^−^*^3^ substitutions/site/year (95% HPD: 2.63 *×* 10*^−^*^3^–3.31 *×* 10*^−^*^3^), consistent with previous reports [39, 41, 42]. Overall, the slower estimated rates of evolution might be partially explained by the time-dependent rate phenomenon [43] in which the presence of historical samples over longer timescales may reduce the rate estimate of more recent samples. We observed the opposite after running a separate Bayesian inference model that was restricted to our 91 university H3 sequences from the 2022–2023 season, and recorded a higher evolutionary rate of 4.06 *×* 10*^−^*^3^ substitutions/site/year (95% HPD: 3.14 *×* 10*^−^*^3^–5.09 *×* 10*^−^*^3^).

We found several highly prevalent H3 mutations that were noted in other surveillance efforts during the 2022-2023 season ( [44, 45]) including N186 which were present in all of our 91 H3 variants (Table 1) and is also in a known epitope region (Fig. 4 panel B). In addition, we identified mutations G225, N186, and I140K in most of our H3 sequences which have been shown to make the electrostatic surface of the receptor binding domain (RBD) more negatively charged than the vaccine strain [46]. We also note the high prevalence (63%) of N96S which is a defining mutation of 2a.3 and 2a.3a.1 variants and creates a new potential N-glycosylation site in the globular head [47]. For N2, we found one instance of mutation S331R (Fig. S5) which has been showed to result in reduced susceptibility to oseltamivir and zanamivir [33].

We estimated vaccine effectiveness via an H3 epitope model with a range of 0.13–0.48 which overlaps with estimates for that year [48, 49]. The abundance of antigenic drift mutations, in addition to our identification of numerous sequons found within HA1 (globular head) with high glycosylation potential (Table 3) likely contributed to moderate vaccine effectiveness on campus for that season.

### Limitations

We note a number of limitations with this study including our focus on a single influenza season on in one university setting. We also acknowledge the potential of sampling bias to impact our Bayesian inference and epidemiological modeling. We attempted to reduce the impact of this by including representative H3 and N2 sequences from GISAID with consideration for diversity across genomic clades and subclades, collection date, and aggregated sampling location. In addition, we lacked metadata including campus residence halls and programs of study which limits our ability to study localized phylodynamics. Including rich metadata in future studies (with ethics approval) would enable more refined analyses such as risk factors for intra-campus transmission, as seen in Holmes *et al.* [50].

We also note that we used the HANA assay developed by Zhou *et al.* [13] which focuses on HA and NA segments and resulted in our high coverage for those segments (Fig. S3) which is inconsistent with depth estimates for full-genome influenza assays [12]. Thus, because of our limitations in amplification and sequencing depth for internal gene segments, we were unable to to perform additional analysis including evolutionary reassortment.

## Conclusions

A limited number of recent studies have focused on sequencing and genomic epidemiology of seasonal influenza variants on college and university campuses since the COVID-19 pandemic. We performed surveillance on one the largest universities in the United States during the 2022-2023 season and observed stark differences to national trends in clade diversity. We identified a local preference of A(H3N2) 3C.2a1b.2a.2a.3a.1 variants which did not assert its dominance of A(H3N2) on the national and international landscape until the following season. Through Bayesian inference and phylodynamics, we observed the impact of multiple domestic and international introductions on fueling local transmission on campus. Additionally, our use of epidemiologic modeling, epitope-based vaccine effectiveness estimates, and amino acid mutation profiling highlight the importance of antigenic drift in sustaining local influenza transmission dynamics. Taken together, we show the importance of genomic epidemiology in semi-closed, highly-dense university settings and its potential for early insight into hemagglutinin clade dominance on a national scale.

## Data availability

We have deposited our assembled sequences into NCBI GenBank under accession numbers: PQ895943–PQ896387 (non-contiguous) for influenza A and PQ895532–PQ895537 for influenza B. In Table S9, we provide a list of the 503 sequences generated and used in this study.

## Acknowledgments

The research reported in this publication was supported by the National Library of Medicine of the National Institutes of Health under award U01LM013129 to R.U.H., M.S., and A.V. The authors acknowledge the support from the ASU Health Clinic Staff and the Research Computing at Arizona State University for access and utilization of the Sol high–performance computing environment [51]. The authors also acknowledge all authors of GISAID sequences used in this manuscript.

## Supplementary materials

### Supplementary figures

**Fig S1.**
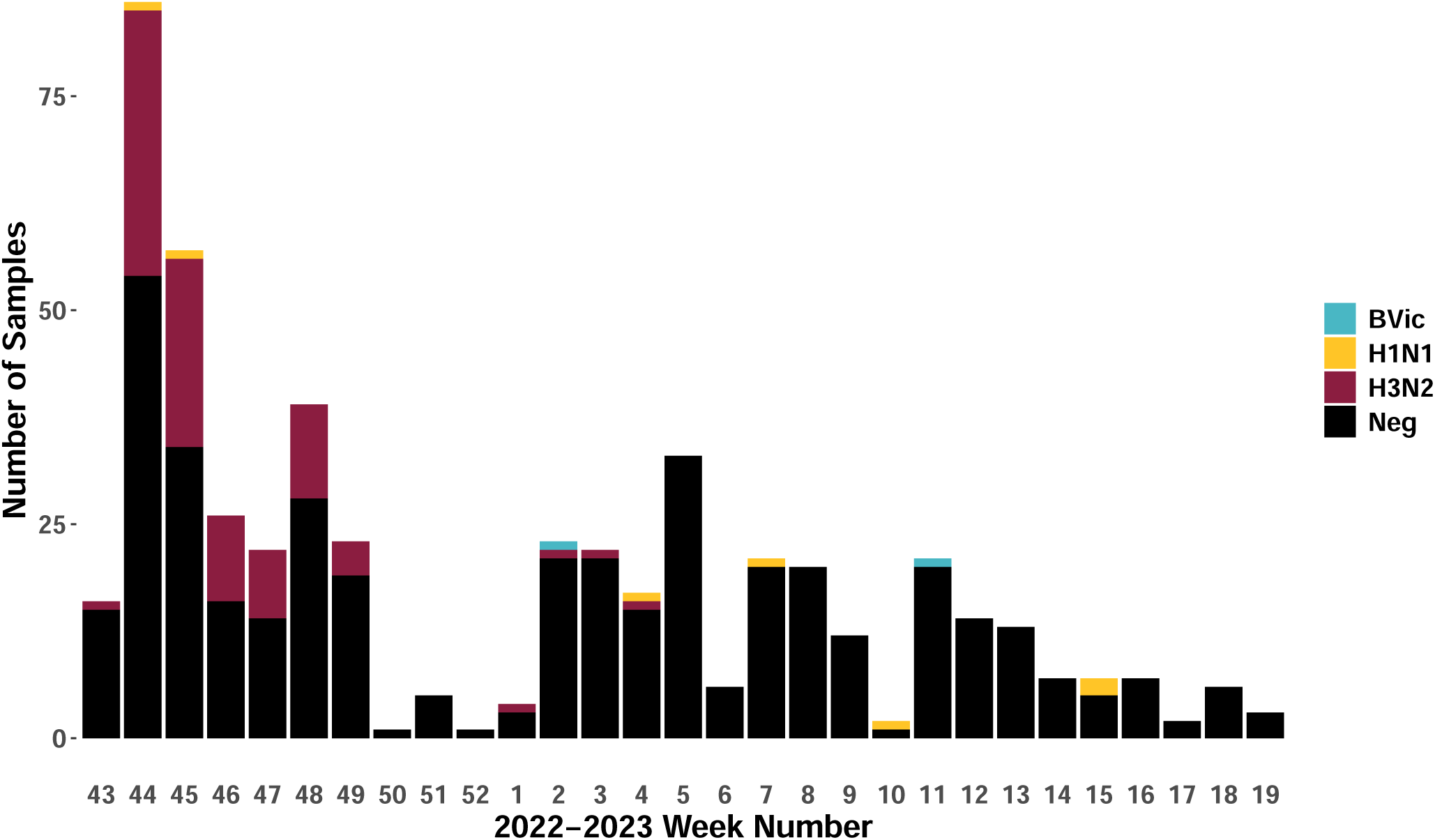
Subtyping results by week at the university health clinic during the 2022–2023 influenza season. Providers tested samples at the student health clinic as part of routine clinical care and labeled as ‘A’, ‘B’, ‘A/B’, or negative via a lateral flow immunoassay. We subtyped positives in our research lab via real–time PCR and pathogen sequencing. We labeled samples as *Undetermined* if our subtype–specific assay yielded an inconclusive result.

**Fig S2.**
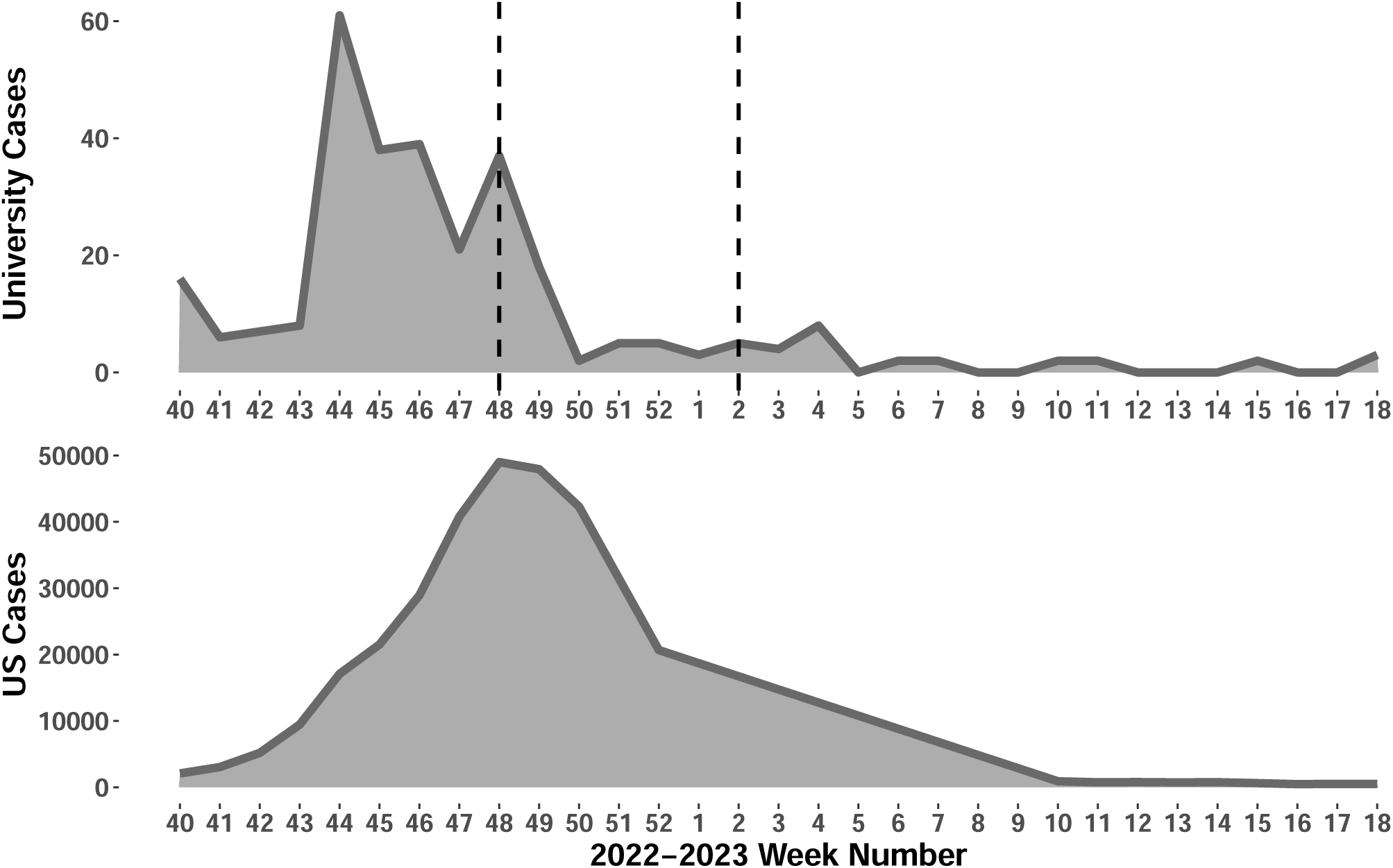
Influenza cases on the university campus (top) and the United States (bottom) during the 2022–2023 season. The two vertical lines mark the end of the fall semester on 2022–12–02 and the start of the spring semester on 2023–01–09, respectively. The US data is from the WHO’s FluNet program [52].

**Fig S3.**
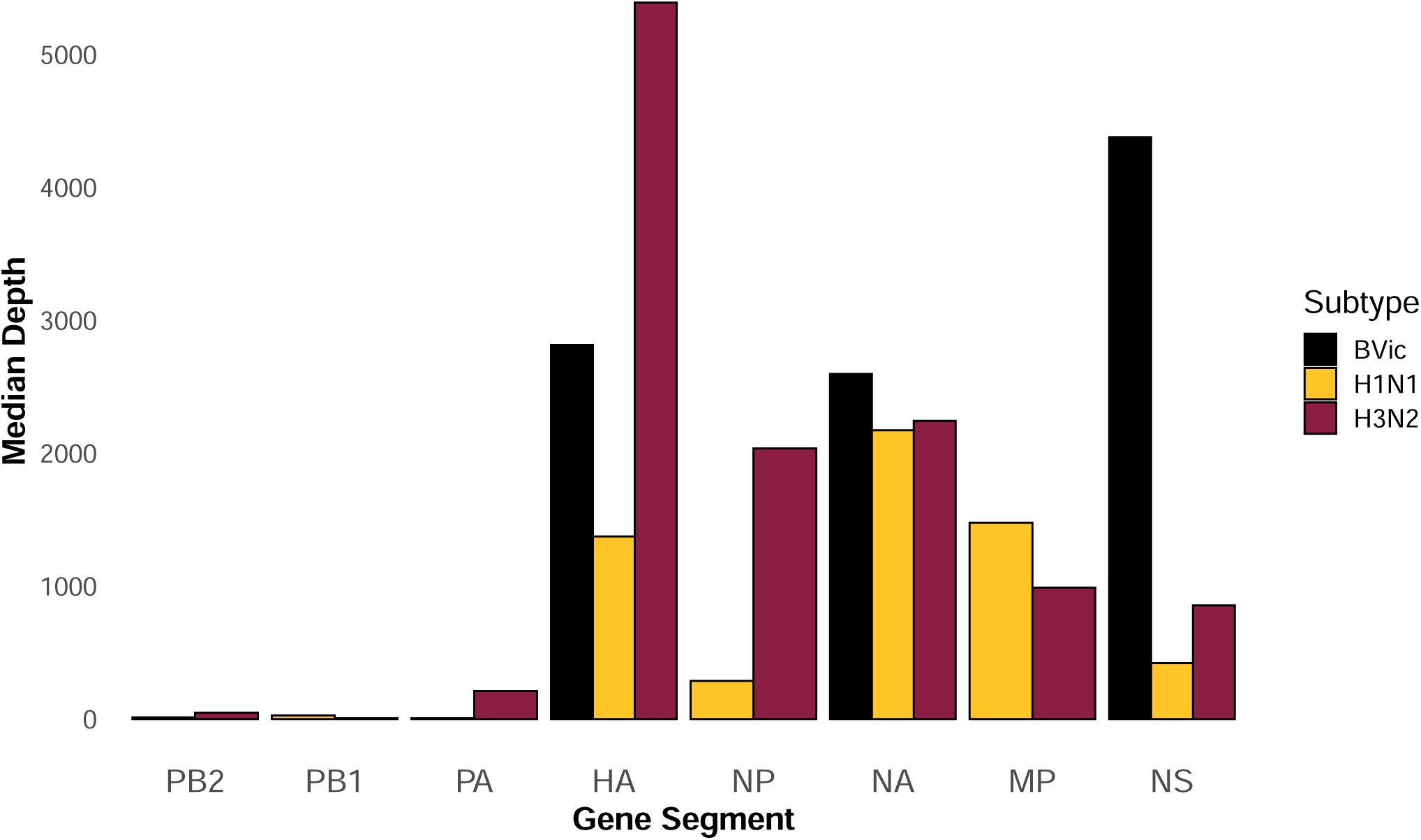
Median coverage depth by subtype and gene segment. The colors indicate the subtype or lineage of influenza: B(Victoria), A(H1N1)pdm09, A(H3N2). Note: While H3 was confirmed in 91 samples, N2 was only confirmed in 76 of these; therefore, it is possible that not all H3 samples were A(H3N2).

**Fig S4.**
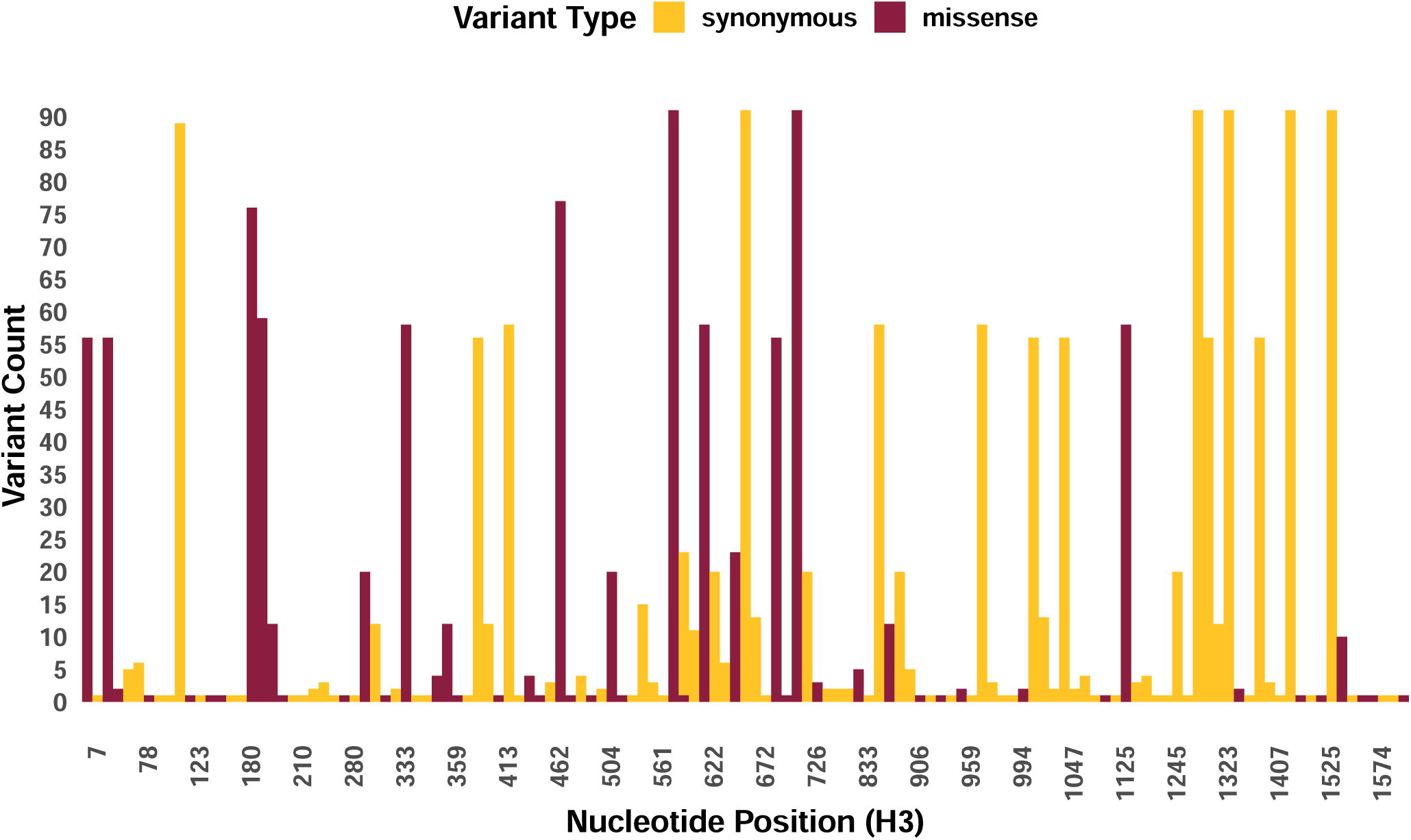
Count of variant types by nucleotide position in the H3 gene segment based on long–read sequencing. Variants were called using A/Darwin/9/2021 as the reference. Colors represent synonymous and missense mutations. We excluded nonsense and other deleterious mutations that are typically purged during influenza virus evolution.

**Fig S5.**
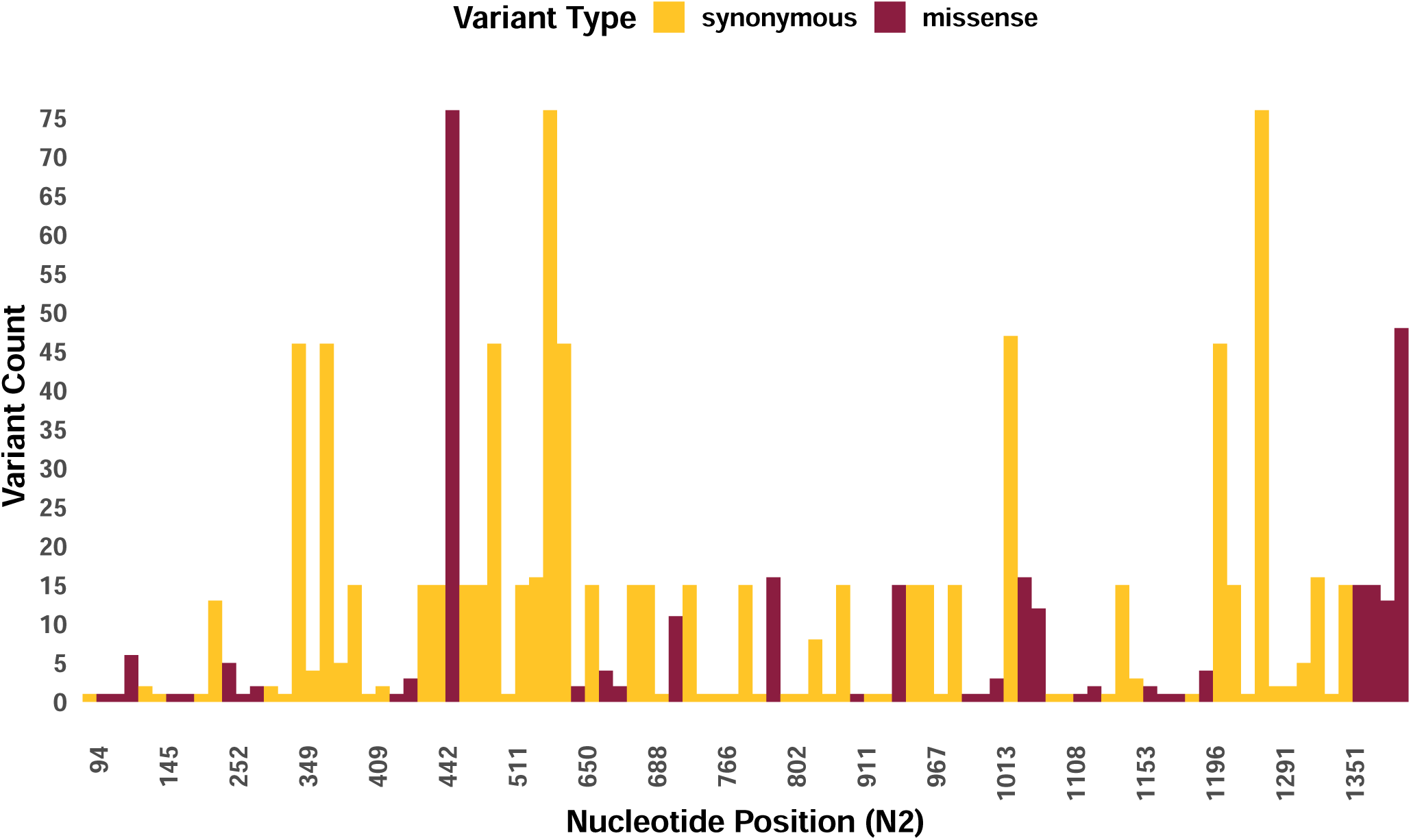
Count of variant types by nucleotide position in the N2 gene segment based on long–read sequencing. Variants were called using A/Darwin/9/2021 as the reference. Colors represent synonymous and missense mutations. We excluded nonsense and other deleterious mutations that are typically purged during influenza virus evolution.

**Fig S6.**
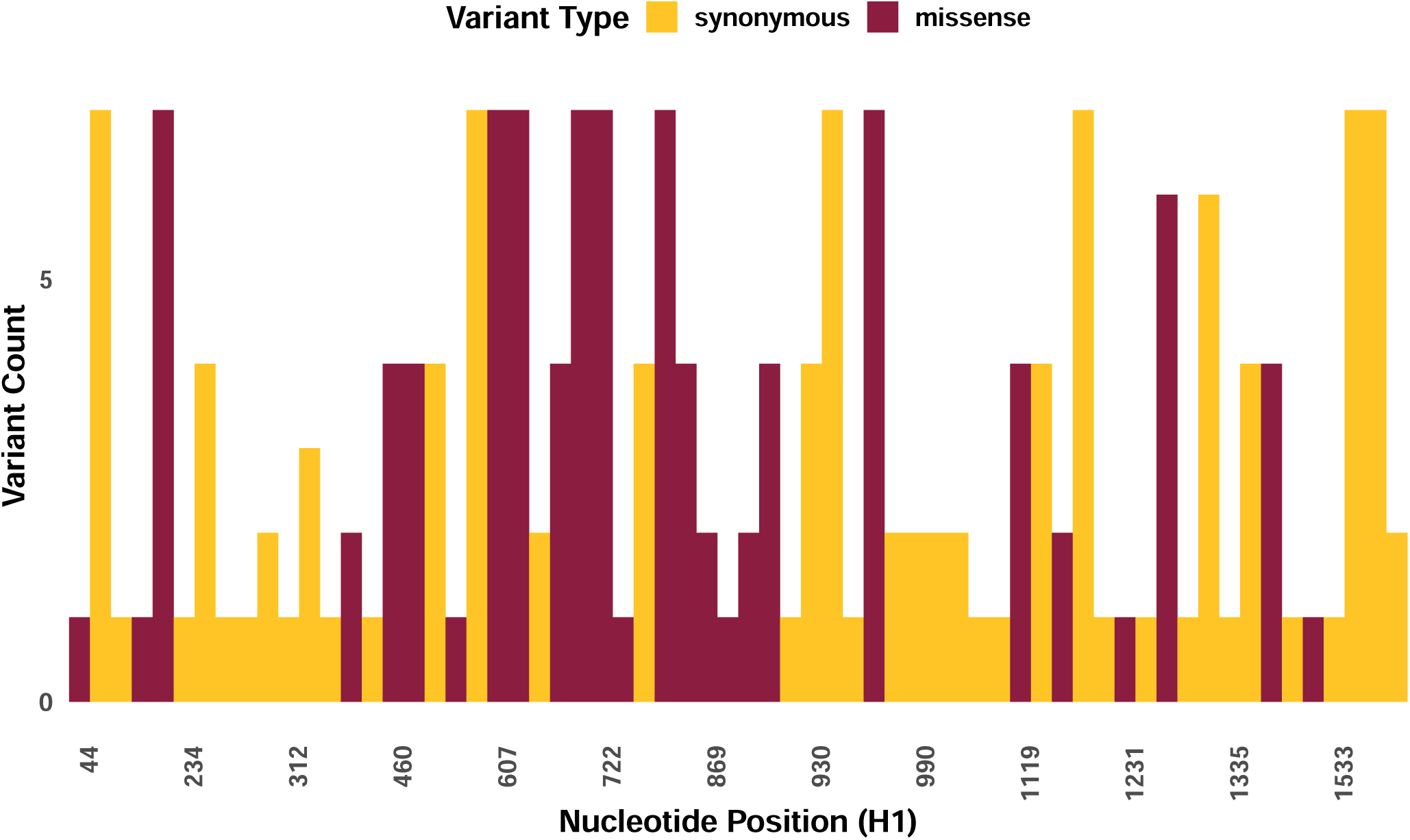
Count of variant type by nucleotide position in the H1 gene segment based on analysis of long–reads. Colors represent synonymous and missense mutations after using A/Victoria/2570/2019 as the reference.

**Fig S7.**
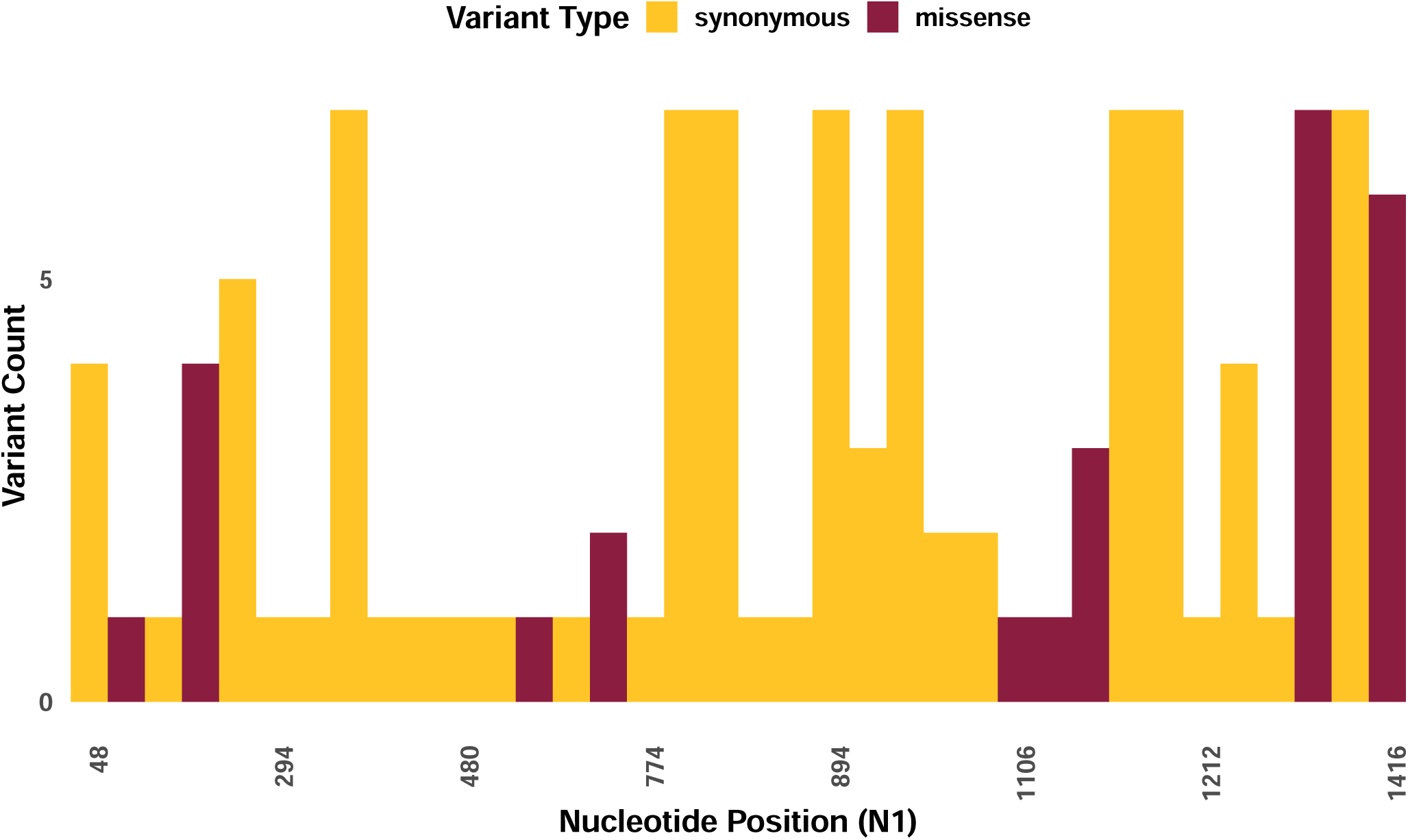
Count of variant type by nucleotide position in the N1 gene segment based on analysis of long–reads. Colors represent synonymous and missense mutations after using A/Victoria/2570/2019 as the reference.

**Fig S8.**
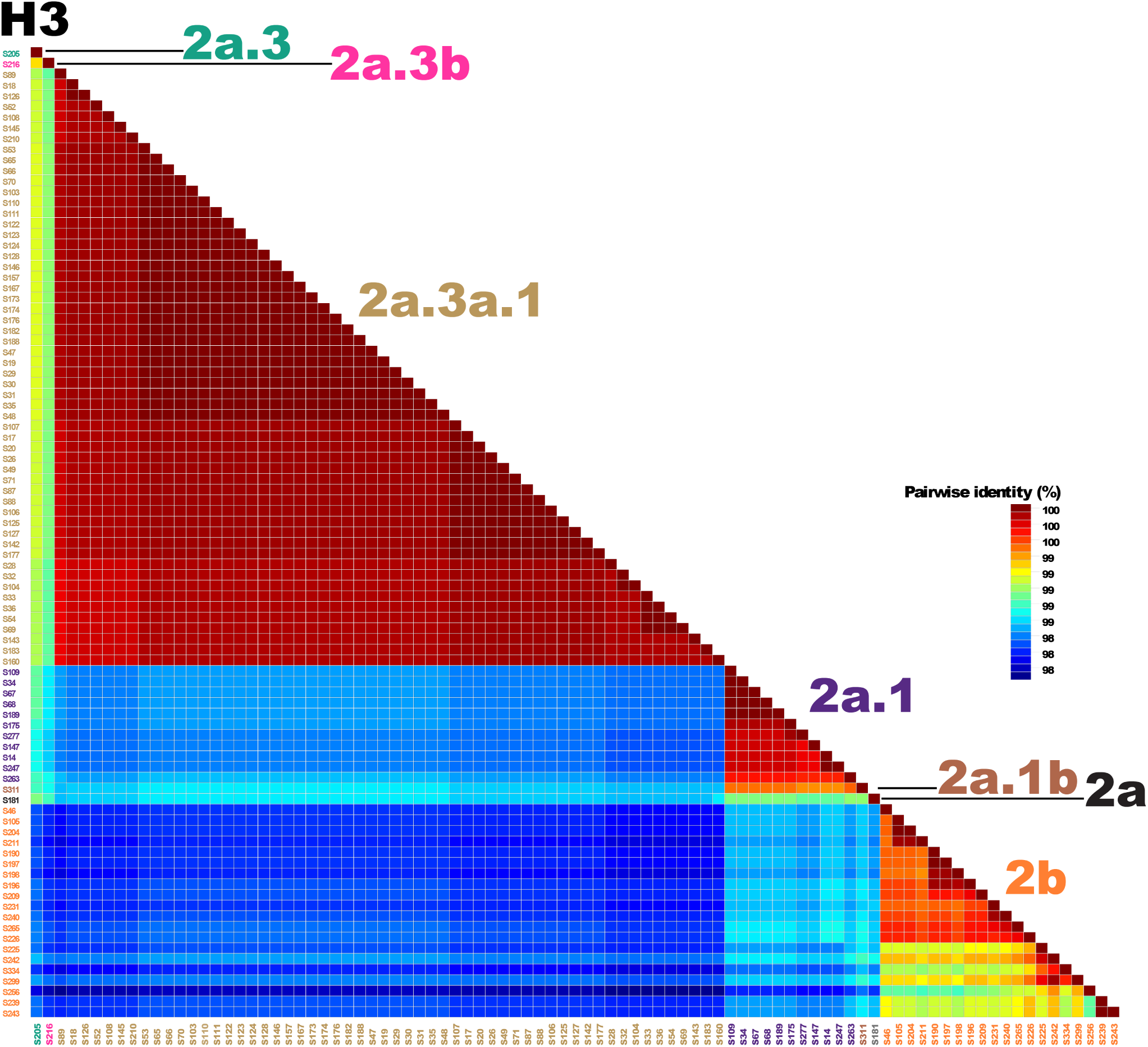
H3 similarity plot via SDT of our 91 sequences. The colors of the sample names reflect their assigned clade as determined by Nextclade. We finalized axis labels and legends in Adobe Illustrator.

**Fig S9.**
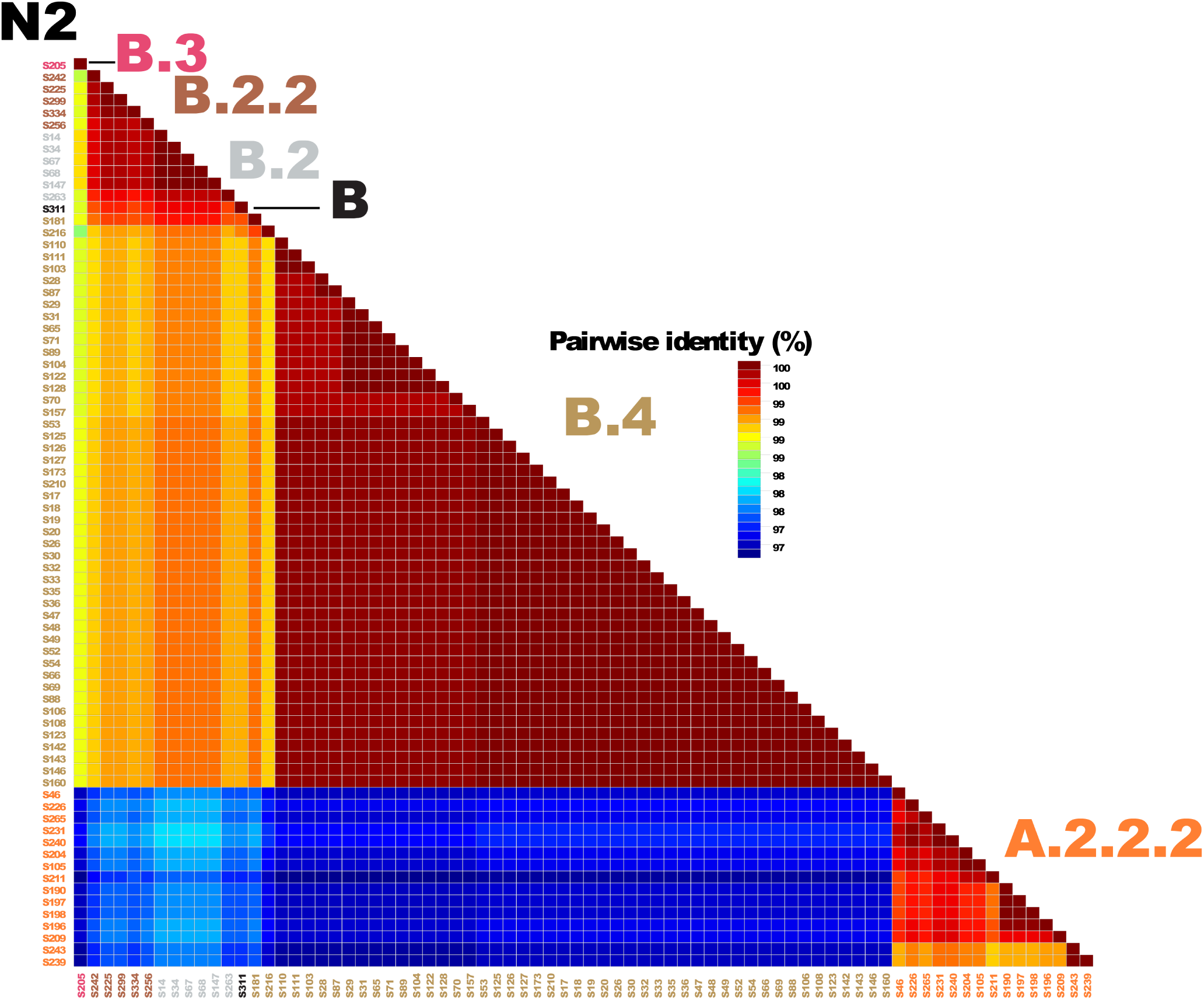
N2 similarity plot via SDT of our 76 sequences. The colors of the sample names reflect their assigned clade as determined by Nextclade. We finalized axis labels and legends in Adobe Illustrator.

**Fig S10.**
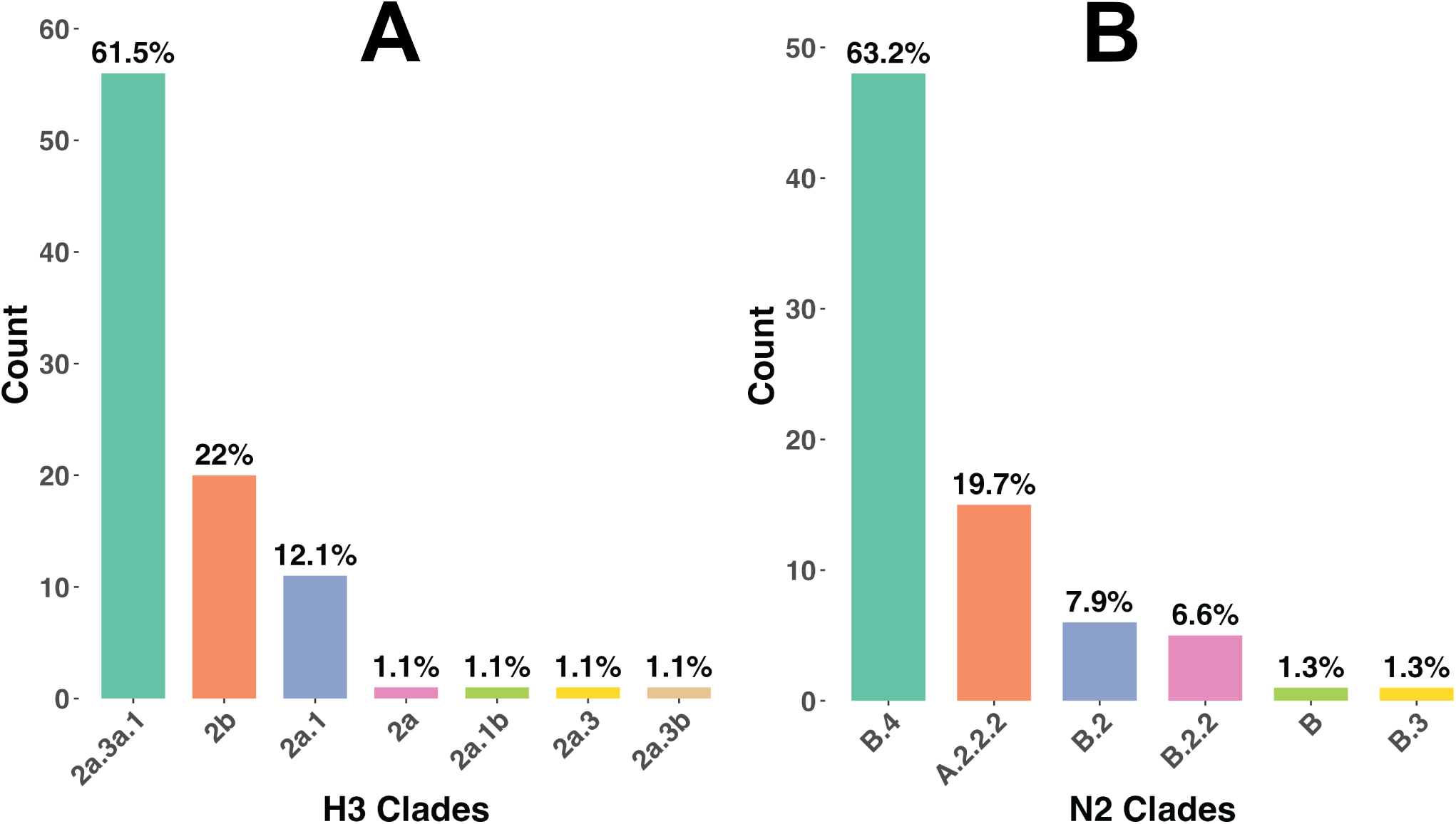
Clade diversity of our H3 and N2 sequences. A) H3 clade distribution for our 91 H3 sequences. B) N2 subclade distribution for our 76 N2 sequences.

**Fig S11.**
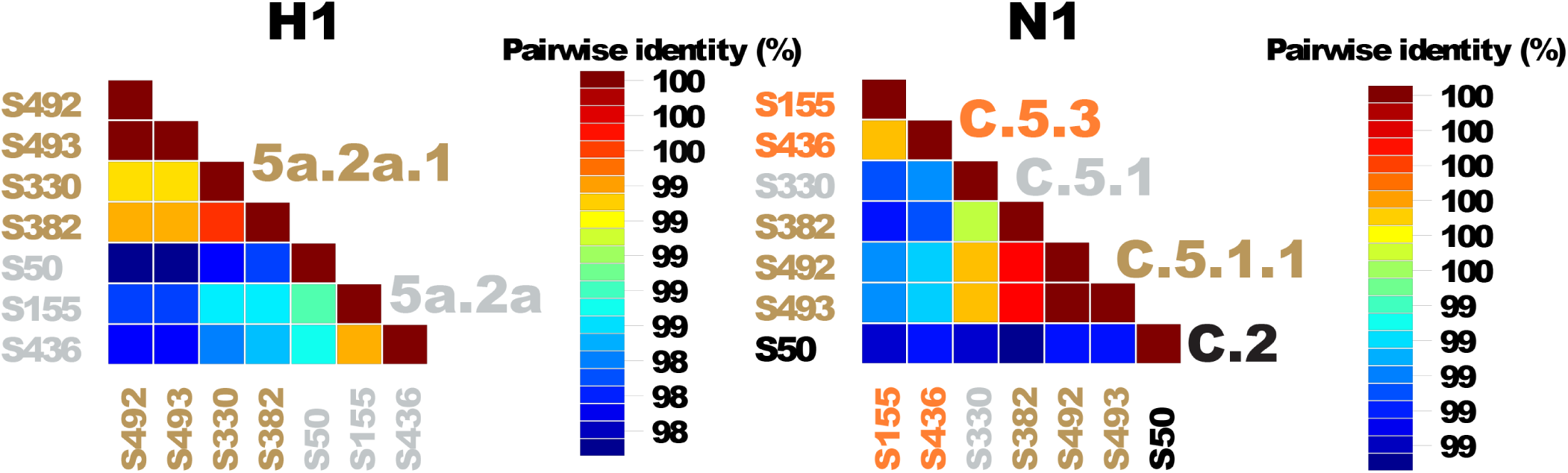
H1 (left) and N1 (right) sequence similarity plots via SDT. The colors of the sample names reflect their assigned clade as determined by Nextclade. We finalized axis labels and legends in Adobe Illustrator.

**Fig S12.**
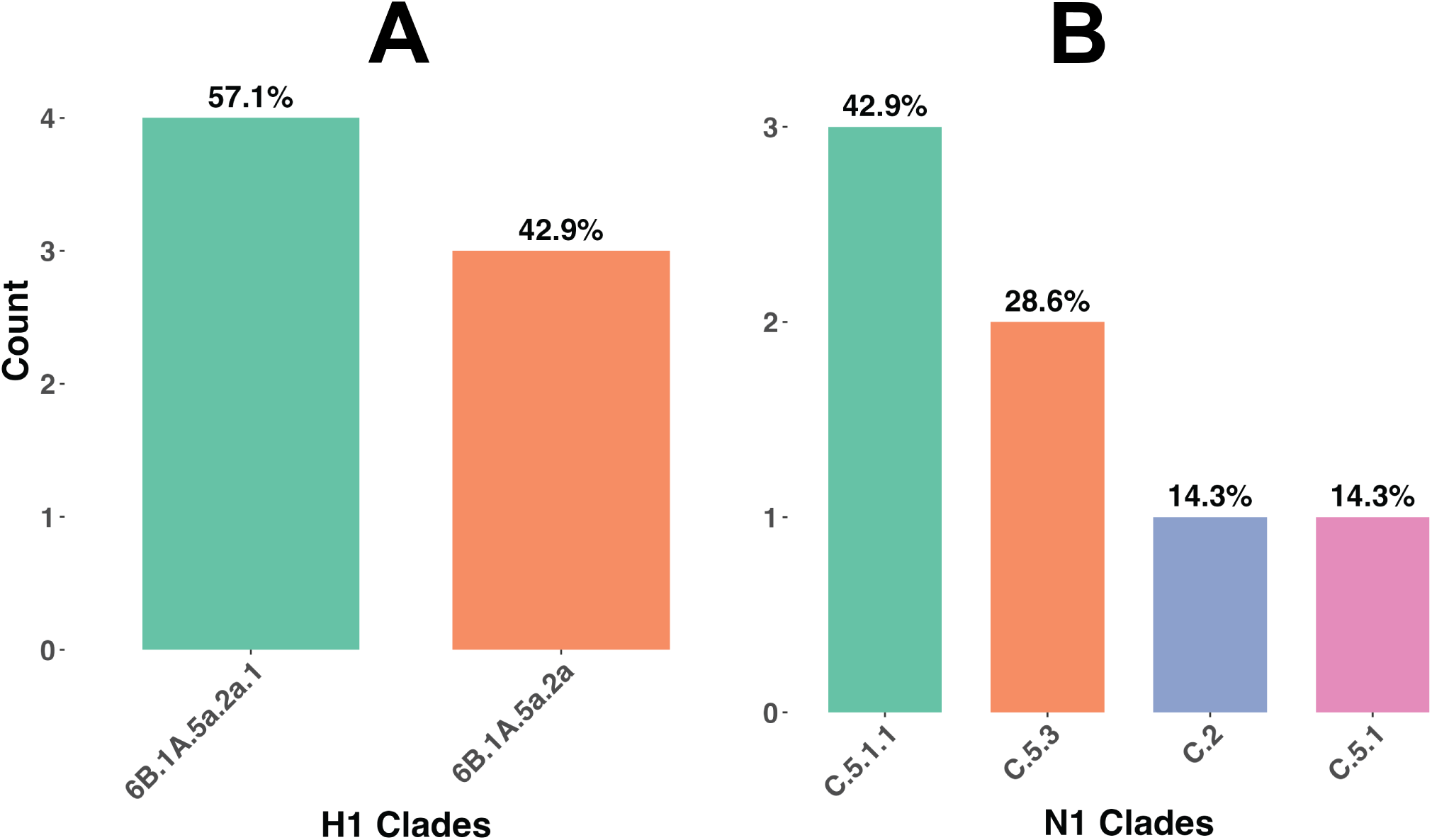
Clade diversity of our H1 and N1 sequences. A) H1 clade distribution for our 7 H1 sequences. B) N1 subclade distribution for our 7 N2 sequences.

**Fig S13.**
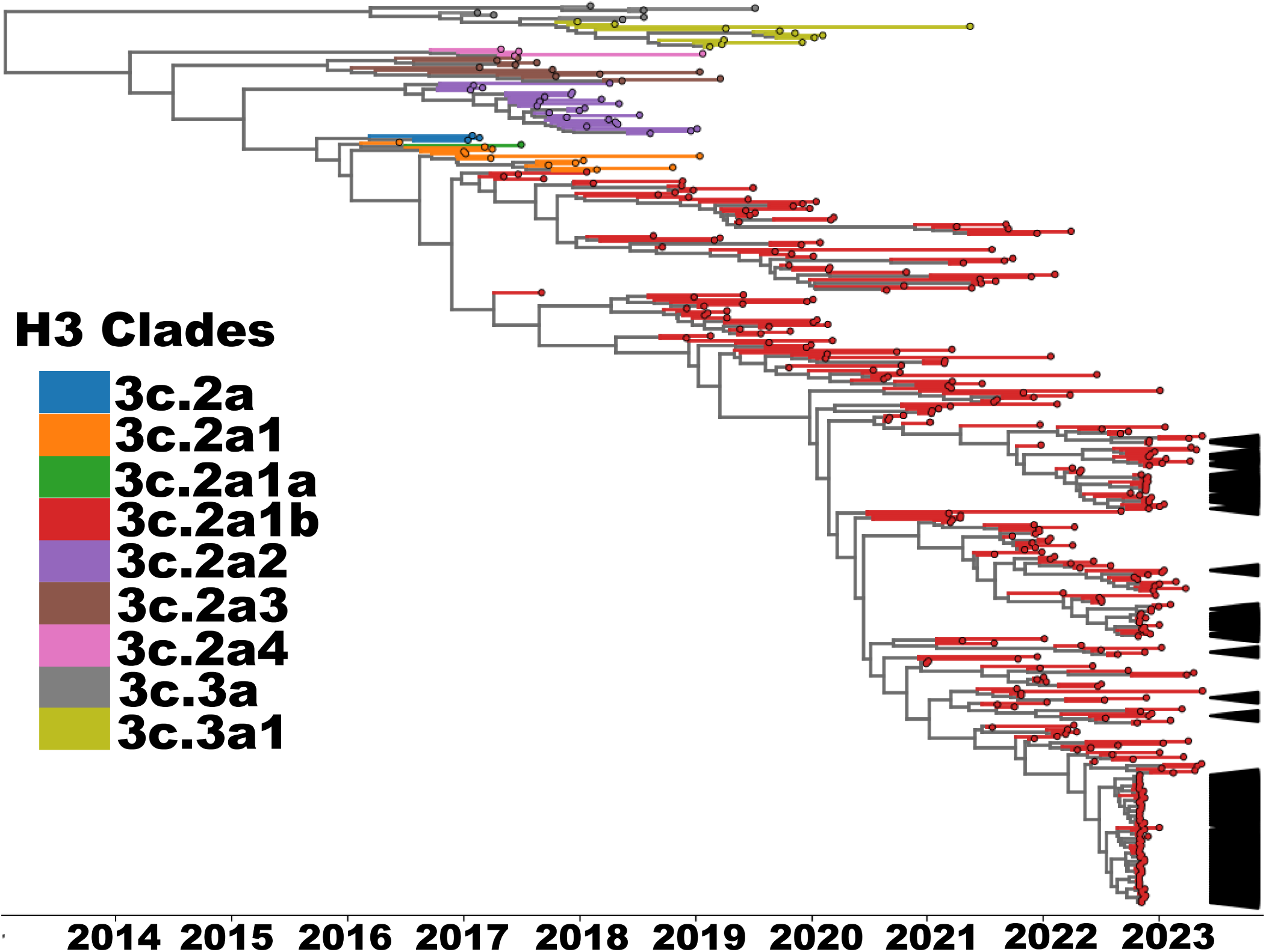
Maximum clade credibility (MCC) phylogeny of H3 hemagglutinin gene sequences collected during the 2022–2023 influenza season. We used BEAST for tree inference and Baltic for visualization and annotation. Tip branches are colored according to major 3C clades (e.g., 3C.2a1, 3C.2a1b, 3C.2a2), which were assigned by mapping each full clade designation to its nearest major clade ancestor for visualization purposes due to the large number of discrete clades. For example, sequences labeled as 3C.2a1b.2a.2a.3a.1 were grouped under *2a1b*. We used arrows to indicate our campus sequences. We finalized axis labels and legends in Adobe Illustrator.

**Fig S14.**
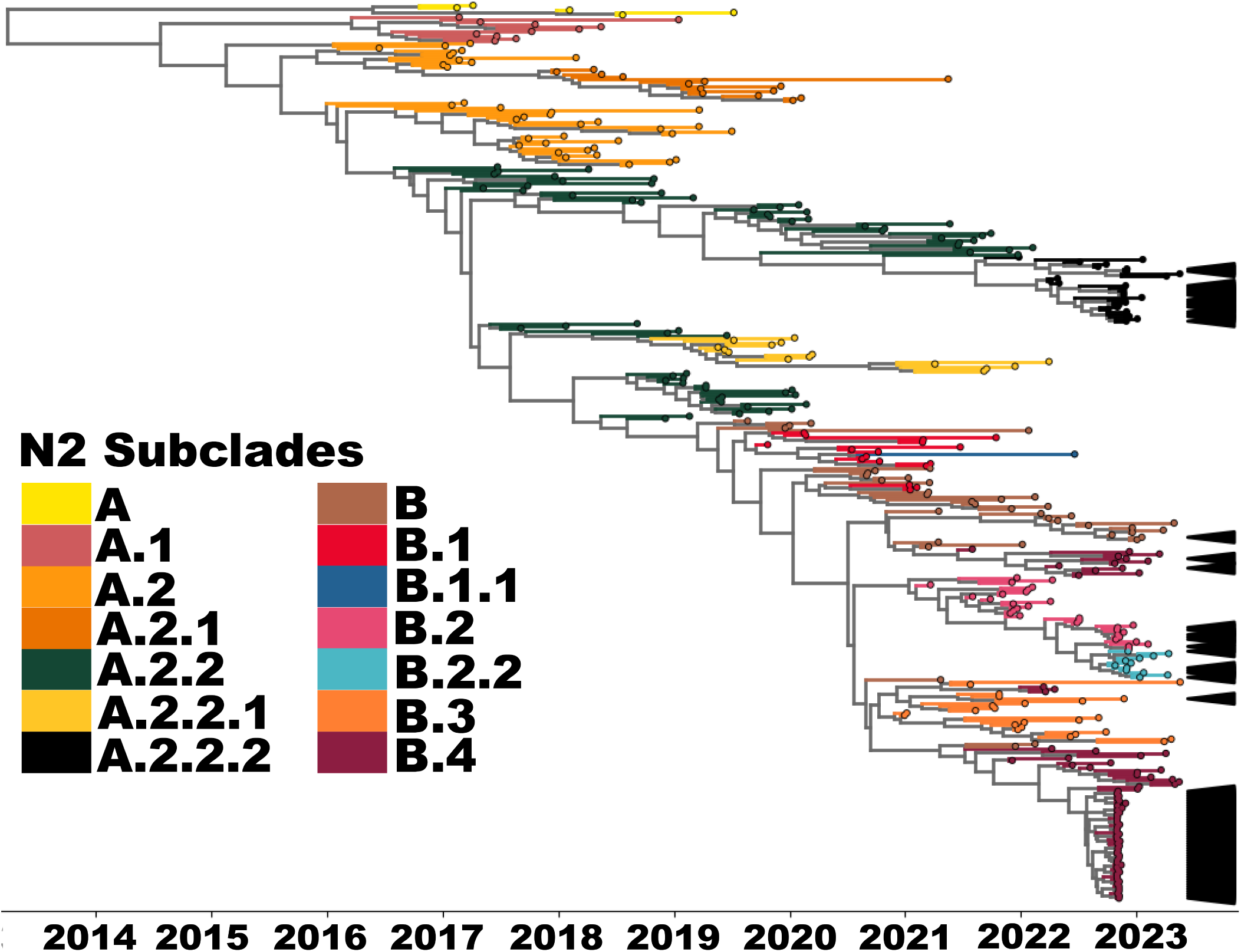
Maximum clade credibility (MCC) phylogeny of N2 neuraminidase gene sequences collected during the 2022–2023 influenza season. We used BEAST for tree inference and Baltic for visualization and annotation. Tip branches are colored according to N2 subclades. We used arrows to indicate our campus sequences. We finalized axis labels and legends in Adobe Illustrator.

**Fig S15.**
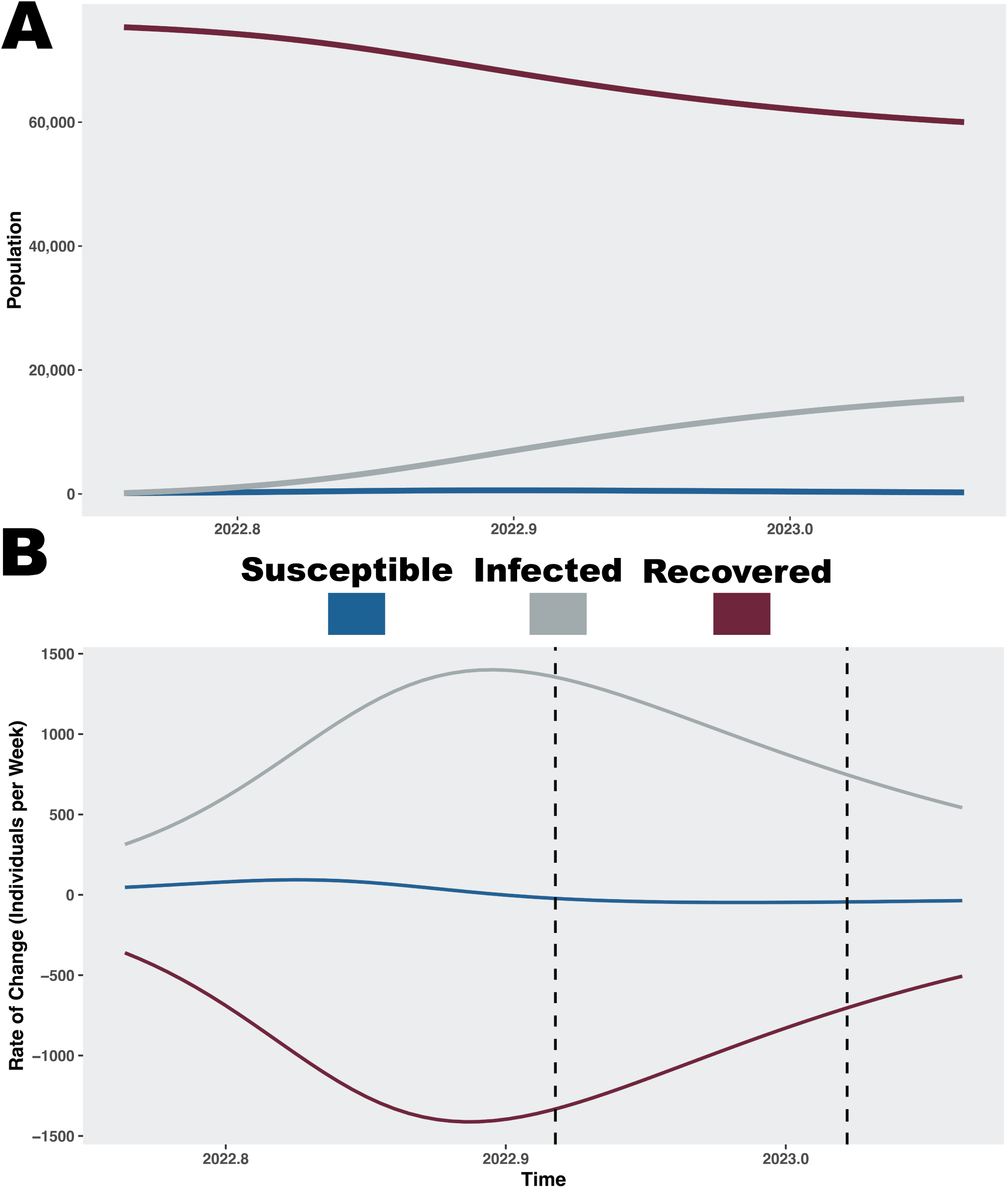
Two–deme SIR model of 2022–2023 H3 influenza on campus. We show the trends of susceptible (S), infected (I), recovered (R) over time (A) and their weekly rate of change (B). The two vertical lines mark the end of the fall semester on 2022–12–02 and the start of the spring semester on 2023–01–09, respectively.

### Supplementary tables

**Table S1.**
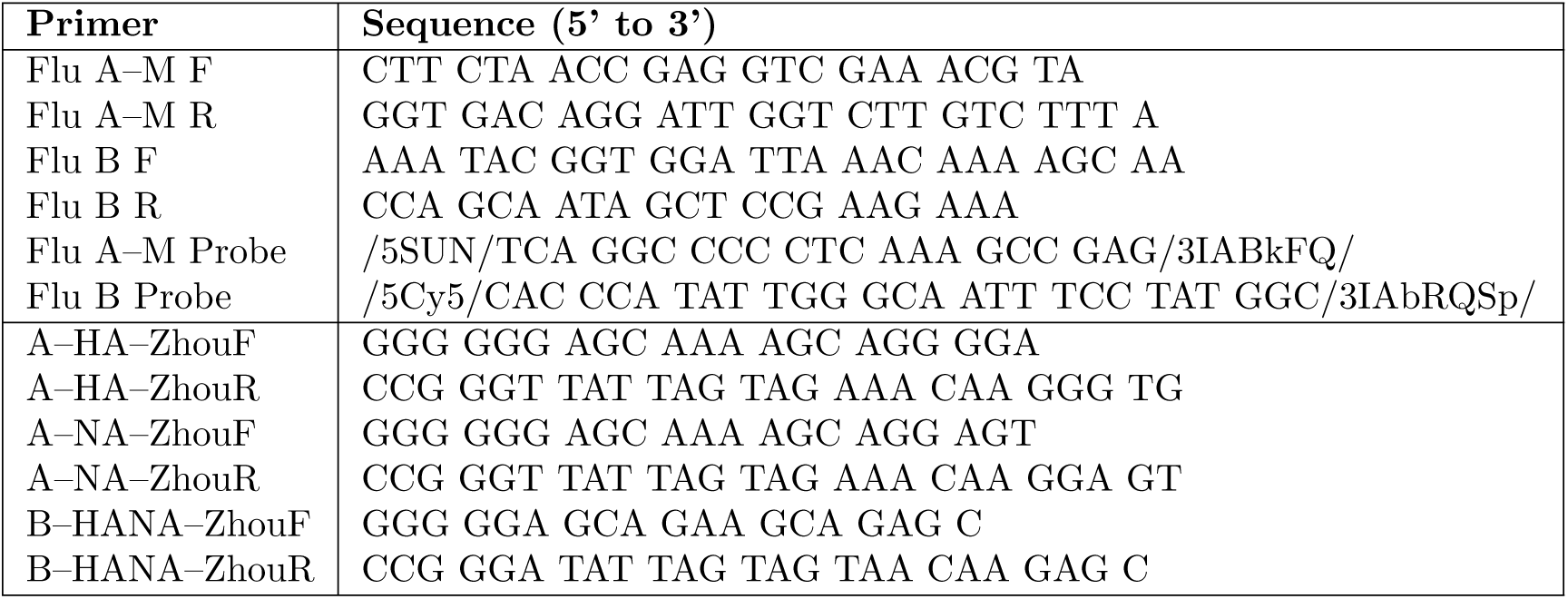
Primers and probes used in the real–time PCR (top) and multiplex PCR assay (bottom) for Influenza A and B detection and amplification.

**Table S2.** 2022–2023 northern hemisphere vaccine reference sequences used in the study.

| Subtype | Strain | GISAI ID |
| --- | --- | --- |
| A(H3N2) | A/Darwin/9/2021 | EPI_ISL_2233240 |
| A(H1N1)pdm09 | A/Victoria/2570/2019 | EPI_ISL_417210 |
| B(Victoria) | B/Austria/1359417/2021 | EPI_ISL_1519459 |

**Table S3.** Long–read sequences (N = 503) that achieved the median coverage threshold of 100, by influenza gene segment.

| Influenza virus | Gene Segment | Number of sequences |
| --- | --- | --- |
| A(H3N2) | PB2 | 30 |
| A(H3N2) | PB1 | 1 |
| A(H3N2) | PA | 55 |
| A(H3N2) | HA | 91 |
| A(H3N2) | NP | 81 |
| A(H3N2) | NA | 76 |
| A(H3N2) | MP | 74 |
| A(H3N2) | NS | 61 |
| A(H1N1)pdm09 | HA | 7 |
| A(H1N1)pdm09 | NP | 4 |
| A(H1N1)pdm09 | NA | 7 |
| A(H1N1)pdm09 | MP | 4 |
| A(H1N1)pdm09 | NS | 6 |
| B(Victoria) | HA | 2 |
| B(Victoria) | NA | 2 |
| B(Victoria) | NS | 2 |

**Table S4.** Log marginal likelihood estimates via stepping–stone and path–sampling for the H3 and N2 Bayesian inference models. We considered different clock models such as strict or uncorrelated lognormal (UCLN) relaxed and tree priors such as constant growth or the nonparametric Bayesian skygrid. The estimates favor the Strict–skygrid combination for both datasets (shaded in gray).

| Data set | Clock Model | Prior | Stepping-stone | Path-sampling |
| --- | --- | --- | --- | --- |
| A/H3 | UCLN | Skygrid | -13,911 | -13,901 |
| A/H3 | Strict | Skygrid | -13,900 | -13,891 |
| A/H3 | UCLN | Constant | -13,944 | -13,937 |
| A/H3 | Strict | Constant | -13,936 | -13,930 |
| A/N2 | UCLN | Skygrid | -10,718 | -10,707 |
| A/N2 | Strict | Skygrid | -10,706 | -10,700 |
| A/N2 | UCLN | Constant | -10,739 | -10,731 |
| A/N2 | Strict | Constant | -10,727 | -10,720 |

**Table S5.**
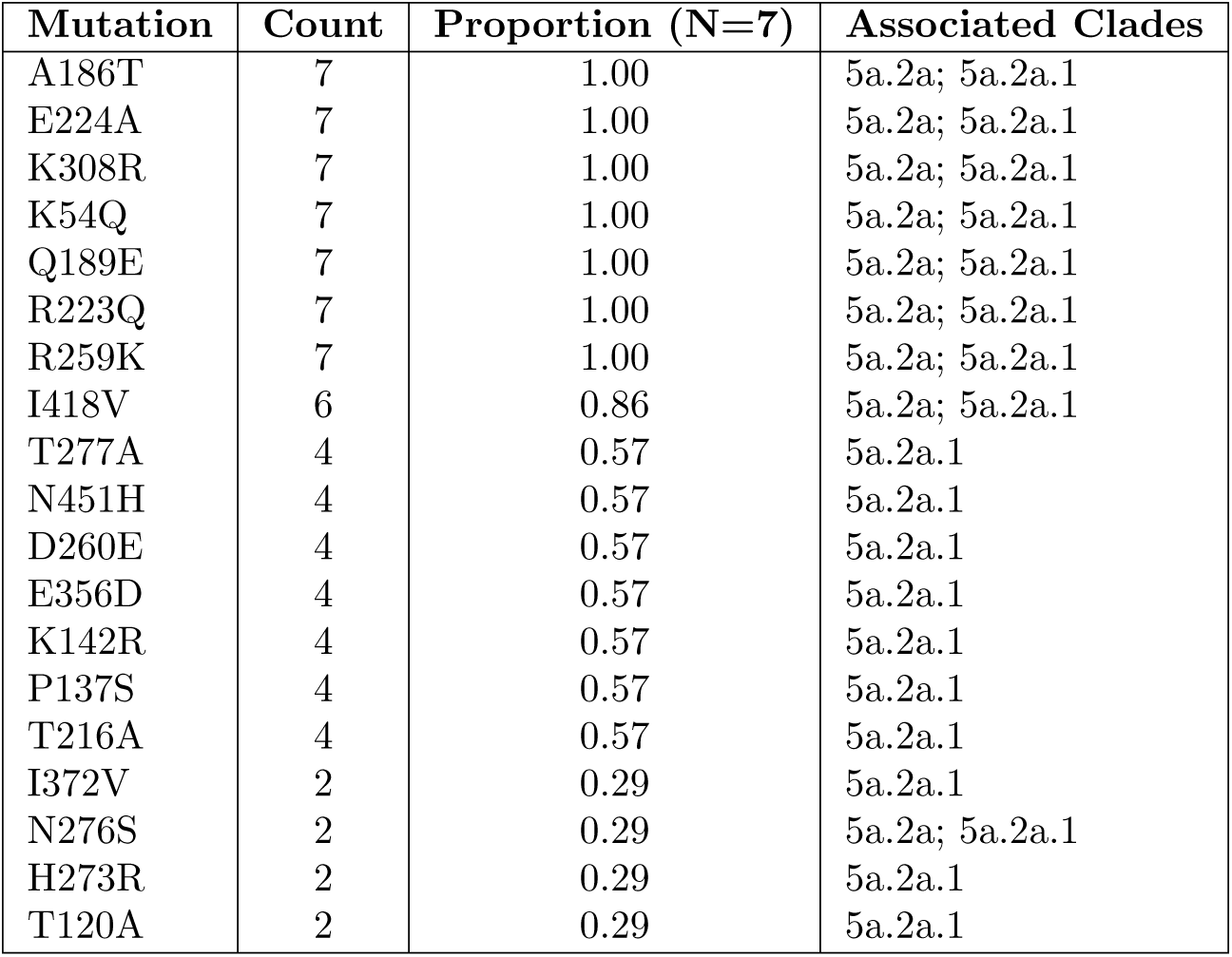
H1 amino acid mutations with a prevalence of *≥* 0.2. For each mutation (H1), we show its count, proportion across our 7 variants, and associated clades (short name).

**Table S6.** N1 amino acid mutations with a prevalence of ≥ 0.2. For each mutation (N1), we show its count, proportion across our 7 variants, and associated clades (short name).

| Mutation | Count | Proportion (N=7) |
| --- | --- | --- |
| V453M | 7 | 1.00 |
| N50D | 4 | 0.57 |
| G382E | 3 | 0.43 |
| S200N | 2 | 0.29 |

**Table S7.** H3 vaccine effectiveness data with dominant epitopes and mutations for 25 unique amino acid sequences. We sorted the table by predicted VE (highest to lowest).

| pEpitope | Dominant Epitope | VE | Epitopes |  |  |  |  |
| --- | --- | --- | --- | --- | --- | --- | --- |
|  |  |  | A | B | C | D | E |
| 0.052 | A | 0.48 | S144G | N186D | D53N | none | none |
| 0.074 | C | 0.41 | none | N186D | D53G,<br>K276R | none | none |
| 0.074 | C | 0.41 | none | N186D | D53G,<br>K276R | none | none |
| 0.074 | C | 0.41 | none | N186D | D53G,<br>K276R | none | none |
| 0.074 | C | 0.41 | I140K | N186D | D53G,<br>K276R | none | none |
| 0.0952 | B | 0.34 | I140K | N186D,<br>S156H | E50K | none | none |
| 0.0952 | B | 0.34 | I140K | N186D,<br>I192F | E50K, D53N | N96S | none |
| 0.0952 | B | 0.34 | I140K | N186D,<br>I192F | E50K, D53N | N96S | none |
| 0.0952 | B | 0.34 | I140K | N186D,<br>I192F | E50K, D53N | N96S | none |
| 0.0952 | B | 0.34 | none | N186D,<br>I192F | D53N | N96S | none |
| 0.0952 | B | 0.34 | I140K | N186D,<br>I192F | E50K, D53N | N96S | none |
| 0.0952 | B | 0.34 | I140K | N186D,<br>S156H | E50K | none | none |
| 0.0952 | B | 0.34 | I140M | N186D,<br>I192F | D53N | N96I | G78S |
| 0.0952 | B | 0.34 | I140K | N186D,<br>S156H | E50K | none | none |
| 0.0952 | B | 0.34 | I140K | N186D,<br>S156H | E50K,<br>A304D | I242M | none |
| 0.0952 | B | 0.34 | I140K | N186D,<br>S156H | E50K | none | S91N, S26N |
| 0.0952 | B | 0.34 | I140K | N186D,<br>I192F | E50K, D53N | N96S | none |
| 0.0952 | B | 0.34 | I140K | N186D,<br>S156H | E50K, S54N | I242M | none |
| 0.0952 | B | 0.34 | I140K | N186D,<br>I192F | E50K, D53N | N96S | none |
| 0.105 | A | 0.31 | I140K,<br>T135A | N186D,<br>S156H | E50K | none | S262N |
| 0.105 | A | 0.31 | S137A,<br>I140K | N186D,<br>S156H | E50K | none | none |
| 0.105 | A | 0.31 | T135A,<br>I140K | N186D,<br>S156H | E50K | none | S262N |
| 0.143 | B | 0.18 | I140K | N186D,<br>L157I, I192F | E50K, D53N | N96S | none |
| 0.158 | A | 0.13 | N122S,<br>T135A,<br>I140K | S156H,<br>N186D | E50K | none | S262N |

**Table S8.** ANOVA results for predicted VE by dominant epitope.

| Source | Sum of Squares | df | Mean Square | F | Sig. |
| --- | --- | --- | --- | --- | --- |
| Between Groups | 305.04 | 2 | 152.52 | 3.00 | .071 |
| Within Groups | 1119.18 | 22 | 50.87 |  |  |
| Total | 1424.22 | 24 |  |  |  |

**Table S9.** GenBank submissions (N = 503) used in this study.

| Strain | PB2 | PB1 | PA | HA | NP | NA | MP | NS |
| --- | --- | --- | --- | --- | --- | --- | --- | --- |
| A/Tempe/S50/2022(H1N1) |  |  |  | PQ895943 | PQ895961 | PQ895960 | PQ895950 | PQ895965 |
| A/Tempe/S155/2022(H1N1) |  |  |  | PQ895944 | PQ895962 | PQ895958 | PQ895951 | PQ895966 |
| A/Tempe/S330/2023(H1N1) |  |  |  | PQ895945 | PQ895963 | PQ895957 |  | PQ895967 |
| A/Tempe/S436/2023(H1N1) |  |  |  | PQ895947 |  | PQ895959 |  | PQ895968 |
| A/Tempe/S492/2023(H1N1) |  |  |  | PQ895948 |  | PQ895955 | PQ895952 | PQ895969 |
| A/Tempe/S493/2023(H1N1) |  |  |  | PQ895949 | PQ895964 | PQ895954 | PQ895953 | PQ895970 |
| A/Tempe/S14/2022(H3N2) | PQ896430 |  | PQ896369 | PQ895971 | PQ896219 | PQ896139 | PQ896061 | PQ896303 |
| A/Tempe/S17/2022(H3N2) |  |  | PQ896370 | PQ895972 | PQ896220 | PQ896140 | PQ896062 | PQ896304 |
| A/Tempe/S18/2022(H3N2) |  |  | PQ896371 | PQ895973 | PQ896221 | PQ896141 | PQ896063 | PQ896305 |
| A/Tempe/S19/2022(H3N2) | PQ896432 |  | PQ896372 | PQ895974 | PQ896222 | PQ896142 | PQ896064 | PQ896306 |
| A/Tempe/S20/2022(H3N2) | PQ896433 |  | PQ896373 | PQ895975 | PQ896223 | PQ896143 | PQ896065 | PQ896307 |
| A/Tempe/S28/2022(H3N2) |  |  |  | PQ895977 | PQ896225 | PQ896145 | PQ896067 | PQ896308 |
| A/Tempe/S29/2022(H3N2) | PQ896434 |  | PQ896374 | PQ895978 | PQ896226 | PQ896146 | PQ896068 | PQ896309 |
| A/Tempe/S30/2022(H3N2) |  |  | PQ896375 | PQ895979 | PQ896227 | PQ896147 | PQ896069 | PQ896310 |
| A/Tempe/S31/2022(H3N2) | PQ896436 |  | PQ896376 | PQ895980 | PQ896228 | PQ896148 | PQ896070 | PQ896311 |
| A/Tempe/S32/2022(H3N2) | PQ896437 |  | PQ896377 | PQ895981 | PQ896229 | PQ896149 | PQ896071 | PQ896312 |
| A/Tempe/S33/2022(H3N2) | PQ896438 |  | PQ896378 | PQ895982 | PQ896230 | PQ896150 | PQ896072 | PQ896313 |
| A/Tempe/S34/2022(H3N2) | PQ896439 |  | PQ896379 | PQ895983 | PQ896231 | PQ896151 | PQ896073 | PQ896314 |
| A/Tempe/S35/2022(H3N2) |  |  | PQ896380 | PQ895984 | PQ896232 | PQ896152 | PQ896074 | PQ896315 |
| A/Tempe/S36/2022(H3N2) |  |  | PQ896381 | PQ895985 | PQ896233 | PQ896153 | PQ896075 | PQ896316 |
| A/Tempe/S46/2022(H3N2) | PQ896440 |  | PQ896382 | PQ895986 | PQ896234 | PQ896154 | PQ896076 | PQ896317 |
| A/Tempe/S47/2022(H3N2) |  |  | PQ896383 | PQ895987 | PQ896235 | PQ896155 | PQ896077 | PQ896318 |
| A/Tempe/S49/2022(H3N2) | PQ896441 |  | PQ896384 | PQ895989 | PQ896237 | PQ896157 | PQ896079 | PQ896319 |
| A/Tempe/S52/2022(H3N2) |  |  | PQ896385 | PQ895990 | PQ896238 | PQ896158 | PQ896080 | PQ896320 |
| A/Tempe/S53/2022(H3N2) |  |  | PQ896386 | PQ895991 | PQ896239 | PQ896159 |  | PQ896321 |
| A/Tempe/S54/2022(H3N2) | PQ896442 |  |  | PQ895992 | PQ896240 | PQ896160 | PQ896082 | PQ896322 |
| A/Tempe/S65/2022(H3N2) |  |  | PQ896387 | PQ895993 | PQ896241 | PQ896161 | PQ896083 | PQ896323 |
| A/Tempe/S67/2022(H3N2) |  |  | PQ896389 | PQ895995 | PQ896243 | PQ896163 | PQ896085 | PQ896324 |
| A/Tempe/S68/2022(H3N2) | PQ896443 |  | PQ896390 | PQ895996 | PQ896244 | PQ896164 | PQ896086 | PQ896325 |
| A/Tempe/S69/2022(H3N2) |  |  | PQ896391 | PQ895997 | PQ896245 | PQ896165 | PQ896087 | PQ896326 |
| A/Tempe/S70/2022(H3N2) |  |  | PQ896392 | PQ895998 | PQ896246 | PQ896166 | PQ896088 | PQ896327 |
| A/Tempe/S87/2022(H3N2) | PQ896444 |  | PQ896394 | PQ896000 | PQ896248 | PQ896168 | PQ896090 | PQ896329 |
| A/Tempe/S88/2022(H3N2) |  |  | PQ896395 | PQ896001 | PQ896249 | PQ896169 | PQ896091 | PQ896330 |
| A/Tempe/S89/2022(H3N2) |  |  | PQ896396 | PQ896002 | PQ896250 | PQ896170 | PQ896092 | PQ896331 |
| A/Tempe/S104/2022(H3N2) |  |  |  | PQ896004 | PQ896252 | PQ896172 | PQ896094 | PQ896333 |
| A/Tempe/S106/2022(H3N2) |  |  |  | PQ896006 | PQ896254 | PQ896174 | PQ896096 | PQ896334 |
| A/Tempe/S111/2022(H3N2) |  |  |  | PQ896011 | PQ896257 | PQ896177 | PQ896098 | PQ896335 |
| A/Tempe/S122/2022(H3N2) | PQ896445 |  | PQ896400 | PQ896012 | PQ896258 | PQ896178 | PQ896099 | PQ896336 |
| A/Tempe/S123/2022(H3N2) | PQ896446 |  | PQ896401 | PQ896013 | PQ896259 | PQ896179 | PQ896100 | PQ896337 |
| A/Tempe/S125/2022(H3N2) |  |  |  | PQ896015 | PQ896261 | PQ896181 | PQ896102 | PQ896339 |
| A/Tempe/S126/2022(H3N2) |  |  |  | PQ896016 | PQ896262 | PQ896182 |  | PQ896340 |
| A/Tempe/S127/2022(H3N2) |  |  | PQ896403 | PQ896017 | PQ896263 | PQ896183 | PQ896104 | PQ896341 |
| A/Tempe/S128/2022(H3N2) |  |  | PQ896404 | PQ896018 | PQ896264 | PQ896184 | PQ896105 | PQ896342 |
| A/Tempe/S142/2022(H3N2) |  |  | PQ896405 | PQ896019 | PQ896265 | PQ896185 | PQ896106 | PQ896343 |
| A/Tempe/S146/2022(H3N2) |  |  | PQ896407 | PQ896022 | PQ896268 | PQ896188 | PQ896107 | PQ896346 |
| A/Tempe/S157/2022(H3N2) |  |  |  | PQ896023 | PQ896269 | PQ896189 | PQ896108 | PQ896347 |
| A/Tempe/S160/2022(H3N2) |  |  |  | PQ896024 | PQ896270 | PQ896190 | PQ896109 | PQ896348 |
| A/Tempe/S183/2022(H3N2) |  |  |  | PQ896033 | PQ896279 |  |  | PQ896350 |
| A/Tempe/S196/2022(H3N2) | PQ896452 |  | PQ896412 | PQ896037 | PQ896283 | PQ896196 | PQ896116 | PQ896351 |
| A/Tempe/S197/2022(H3N2) | PQ896453 |  | PQ896413 | PQ896038 | PQ896284 | PQ896197 | PQ896117 | PQ896352 |
| A/Tempe/S204/2022(H3N2) | PQ896455 |  | PQ896415 | PQ896040 | PQ896286 | PQ896199 | PQ896119 | PQ896353 |
| A/Tempe/S205/2022(H3N2) | PQ896456 |  |  | PQ896041 | PQ896287 | PQ896200 | PQ896120 | PQ896354 |
| A/Tempe/S209/2022(H3N2) |  |  |  | PQ896042 | PQ896288 | PQ896201 | PQ896121 | PQ896355 |
| A/Tempe/S210/2022(H3N2) |  |  |  | PQ896043 | PQ896289 | PQ896202 | PQ896122 | PQ896356 |
| A/Tempe/S211/2022(H3N2) | PQ896458 |  |  | PQ896044 | PQ896290 | PQ896203 | PQ896123 | PQ896357 |
| A/Tempe/S225/2022(H3N2) |  |  |  | PQ896046 | PQ896292 | PQ896205 | PQ896125 | PQ896358 |
| A/Tempe/S226/2022(H3N2) | PQ896459 |  | PQ896420 | PQ896047 | PQ896293 | PQ896206 | PQ896126 | PQ896359 |
| A/Tempe/S240/2022(H3N2) | PQ896460 |  | PQ896421 | PQ896050 | PQ896295 | PQ896209 | PQ896129 | PQ896360 |
| A/Tempe/S242/2022(H3N2) |  |  | PQ896422 | PQ896051 | PQ896296 | PQ896210 | PQ896130 | PQ896361 |
| A/Tempe/S243/2022(H3N2) |  |  |  | PQ896052 | PQ896297 | PQ896211 | PQ896131 | PQ896362 |
| A/Tempe/S256/2022(H3N2) | PQ896462 |  | PQ896423 | PQ896054 | PQ896299 | PQ896213 | PQ896132 | PQ896363 |
| A/Tempe/S263/2022(H3N2) |  |  | PQ896424 | PQ896055 | PQ896300 | PQ896214 | PQ896133 | PQ896364 |
| A/Tempe/S265/2022(H3N2) |  |  | PQ896425 | PQ896056 | PQ896301 | PQ896215 | PQ896134 | PQ896365 |
| A/Tempe/S299/2023(H3N2) |  |  | PQ896427 | PQ896058 |  | PQ896216 | PQ896136 | PQ896366 |
| A/Tempe/S311/2023(H3N2) | PQ896467 |  | PQ896428 | PQ896059 |  | PQ896217 | PQ896137 | PQ896367 |
| A/Tempe/S334/2023(H3N2) | PQ896468 |  | PQ896429 | PQ896060 | PQ896302 | PQ896218 | PQ896138 | PQ896368 |
| A/Tempe/S147/2022(H3N2) | PQ896475 | PQ896476 | PQ896474 | PQ896469 | PQ896472 | PQ896471 | PQ896470 | PQ896473 |
| A/Tempe/S48/2022(H3N2) |  |  |  | PQ895988 | PQ896236 | PQ896156 | PQ896078 |  |
| A/Tempe/S66/2022(H3N2) |  |  |  | PQ895994 | PQ896242 | PQ896162 | PQ896084 |  |
| A/Tempe/S71/2022(H3N2) |  |  | PQ896393 | PQ895999 | PQ896247 | PQ896167 | PQ896089 |  |
| A/Tempe/S103/2022(H3N2) |  |  | PQ896397 | PQ896003 | PQ896251 | PQ896171 | PQ896093 |  |
| A/Tempe/S105/2022(H3N2) |  |  | PQ896399 | PQ896005 | PQ896253 | PQ896173 | PQ896095 |  |
| A/Tempe/S108/2022(H3N2) |  |  |  | PQ896008 | PQ896255 | PQ896175 | PQ896097 |  |
| A/Tempe/S173/2022(H3N2) |  |  |  | PQ896026 | PQ896272 | PQ896192 |  |  |
| A/Tempe/S175/2022(H3N2) | PQ896451 |  | PQ896408 | PQ896028 | PQ896274 |  | PQ896112 |  |
| A/Tempe/S181/2022(H3N2) |  |  |  | PQ896031 | PQ896277 | PQ896193 | PQ896113 |  |
| A/Tempe/S182/2022(H3N2) |  |  |  | PQ896032 | PQ896278 |  | PQ896114 |  |
| A/Tempe/S189/2022(H3N2) |  |  |  | PQ896035 |  |  | PQ896115 |  |
| A/Tempe/S198/2022(H3N2) | PQ896454 |  | PQ896414 | PQ896039 | PQ896285 | PQ896198 | PQ896118 |  |
| A/Tempe/S216/2022(H3N2) |  |  | PQ896418 | PQ896045 | PQ896291 |  | PQ896124 |  |
| A/Tempe/S231/2022(H3N2) |  |  |  | PQ896048 |  | PQ896207 | PQ896127 |  |
| A/Tempe/S239/2022(H3N2) |  |  |  | PQ896049 | PQ896294 | PQ896208 | PQ896128 |  |
| A/Tempe/S277/2023(H3N2) | PQ896465 |  | PQ896426 | PQ896057 |  |  | PQ896135 |  |
| A/Tempe/S382/2023(H1N1) |  |  |  | PQ895946 |  | PQ895956 |  |  |
| A/Tempe/S26/2022(H3N2) |  |  |  | PQ895976 | PQ896224 | PQ896144 |  |  |
| A/Tempe/S110/2022(H3N2) |  |  |  | PQ896010 | PQ896256 | PQ896176 |  |  |
| A/Tempe/S143/2022(H3N2) |  |  | PQ896406 | PQ896020 | PQ896266 | PQ896186 |  |  |
| A/Tempe/S190/2022(H3N2) |  |  | PQ896411 | PQ896036 |  | PQ896195 |  |  |
| A/Tempe/S124/2022(H3N2) |  |  |  | PQ896014 | PQ896260 |  |  |  |
| A/Tempe/S167/2022(H3N2) | PQ896450 |  |  | PQ896025 | PQ896271 |  |  |  |
| A/Tempe/S174/2022(H3N2) |  |  |  | PQ896027 | PQ896273 |  |  |  |
| A/Tempe/S176/2022(H3N2) |  |  |  | PQ896029 | PQ896275 |  |  |  |
| A/Tempe/S177/2022(H3N2) |  |  |  | PQ896030 | PQ896276 |  |  |  |
| A/Tempe/S188/2022(H3N2) |  |  | PQ896410 | PQ896034 | PQ896280 |  |  |  |
| A/Tempe/S107/2022(H3N2) |  |  |  | PQ896007 |  |  |  |  |
| A/Tempe/S109/2022(H3N2) |  |  |  | PQ896009 |  |  |  |  |
| A/Tempe/S145/2022(H3N2) |  |  |  | PQ896021 |  |  |  |  |
| A/Tempe/S247/2022(H3N2) |  |  |  | PQ896053 |  |  |  |  |
| B/Tempe/S281/2023(B) |  |  |  | PQ895533 |  | PQ895535 |  | PQ895536 |
| B/Tempe/S448/2023(B) |  |  |  | PQ895532 |  | PQ895534 |  | PQ895537 |

## Supplemental methodology

### Sequencing, assembly, and variant analysis

We sequenced our long-read libraries on a MinION Mk1b with a flongle R9.4.1 flow cell (Oxford Nanopore, Oxford, UK) and utilized Guppy v6.2.7 (as part of MinKNOW v22.08.4) for basecalling. We used fastqc [53] and nanoplot [54] on galaxy [55] for quality assurance. We trimmed adapters from the raw reads using porechop 0.2.4 [56] and filtered out reads with less than 500 nucleotides and phred scores below 15 using fastplong [57]. We re-checked for QC using fastqc and nanoplot and assembled them via the Centers for Disease Control and Prevention’s (CDC) IRMA pipeline using the *FLU-minion* module [58] and minimap2 for assembly [59]. For our reference set, we used the recommended 2022–2023 northern Hemisphere vaccine sequences (Table S2). For the remaining steps in our pipeline, we used Medaka v1.12.0 for polishing and variant analysis [60], SnpEff for annotation and functional effect prediction [61], and bcftools for querying relevant columns in the variant files [62]. Finally, we used the NCBI’s Influenza Virus Sequence Annotation Tool [63] for subtype confirmation and identification of any remaining errors in the consensus sequence.

### Sequence reference data

To construct a representative dataset of influenza sequences for phylogenetic analysis, we downloaded metadata from GISAID and implemented a structured sub-sampling strategy that included both recent and historical sequences. Although our primary focus was on the 2022–2023 influenza season, we extended our sampling to include sequences collected as far back as the 2016–2017 season. This broader temporal coverage was necessary to place our focal sequences in historical context, to better understand patterns of antigenic drift over time, and to accurately root the phylogeny.

We began by filtering the dataset to retain only sequences with valid collection dates. For recent years (2021–2023), we sampled up to two sequences per month to maintain high temporal resolution during our primary study period. For older years (2017–2020), we sampled one sequence per calendar quarter to provide temporal anchoring points without overwhelming the dataset. To enhance representation of genetic diversity, we selected at least one sequence per clade per season. Additionally, to account for spatial variation, we sampled one representative per region per quarter.

We combined all sampled sequences across these strategies and removed duplicates to finalize our dataset. Sequences selected via any of these criteria were flagged for downstream analyses. In Geneious Prime v2024.0.7 (Dotmatics, Boston, MA USA), we excluded sequences with incomplete coding regions, such as those lacking start and/or stop codons.

### Bayesian phylogenetic inference

We used BEAUti v1.10.4 [20] to specify different population growth priors including a constant growth and a non-parametric Bayesian skygrid coalescent model [64]. We evaluated their fit to the data by calculating the log-marginal likelihood values via stepping-stone and path-sampling analysis [65] which suggested the use of the nonparametric Skyrgid for both H3 and N2 data sets (Table S4). We ran separate Markov-chain Monte Carlo (MCMC) simulations for 5 *x* 10^7^ steps and sampled every 5 *x* 10^3^ steps. We combined posterior data via LogCombiner [20] and checked for convergence via Tracer v1.7.1 [66] including that all effective sampling size (ESS) values were *≥* 200. We generated a maximum clade credibility (MCC) tree via TreeAnnotator v1.10.4 [20] from the combined set of posterior trees.

To understand the spatiotemporal and evolutionary dynamics of influenza H3 clades and N2 subclades circulating at the university, including the number and timing of viral migration events, we selected 1,000 trees from the combined posterior distribution and used them as empirical input for discrete trait ancestral state reconstruction in BEAST v1.10.4 [20]. We annotated each sequence with a discrete location attribute corresponding to its sampling region (e.g., North America, Europe, Asia, university). We applied a discrete trait model with Bayesian stochastic search variable selection (BSSVS) to estimate asymmetric transition rates between locations while incorporating model uncertainty [67]. To infer the geographic sources and timing of introductions into the university, we recorded Markov jumps from all regions to the university [68]. We ran the analysis for 2 *x* 10^6^ steps, sampling every 2 *x* 10^2^ steps. After discarding 10% burn-in, we summarized the posterior tree set using TreeAnnotator v1.10.4 [20] to generate a maximum clade credibility (MCC) tree. From this tree, we extracted the number, timing, and geographic origin of introduction events, along with the number of descendant tips per introduction, using a custom R script.

### Parameters and equations for Bayesian epidemiologic model

*I* = AZ deme

*N_r_*= global deme

*I(t)* = # of people that are infectious with the virus at time *t*

*S(t)* = # of people that are susceptible to H3N2 infection at time *t*

β = *per capita* transmission rate

*R(t)* = # of people that are immune due to previous infection

*N(t)* = virus effective population size

η = migration rate

γ = recovery rate

*R0* = basic reproduction number

*N* = *S* + *I* + *R*;

*λ*(*t ≥* 2022.75) = *βI*(*t*) *× S*(*t*)*/N λ*(*t <* 2022.75) = *γ*

*R0* = *β/γ*

[24]

γ = 4.8 lognormal (informed by [69])

*R0* = 1.1

η = 2.0

*S* = 54,000

*I* = 75

*R* = 0

## Notes

### Competing Interest Statement

The authors have declared no competing interest.

### Author Declarations

Arizona State University Institutional Review Board determined that the activity described in the manuscript is not research involving human subjects as defined by DHHS and FDA regulation.

